# Impact of COVID-19 on mental illness in vaccinated and unvaccinated people: a population-based cohort study in OpenSAFELY

**DOI:** 10.1101/2023.12.06.23299602

**Authors:** Venexia M Walker, Praveetha Patalay, Jose Ignacio Cuitun Coronado, Rachel Denholm, Harriet Forbes, Jean Stafford, Bettina Moltrecht, Tom Palmer, Alex Walker, Ellen J. Thompson, Kurt Taylor, Genevieve Cezard, Elsie M F Horne, Yinghui Wei, Marwa Al Arab, Rochelle Knight, Louis Fisher, Jon Massey, Simon Davy, Amir Mehrkar, Seb Bacon, Ben Goldacre, Angela Wood, Nishi Chaturvedi, John Macleod, Ann John, Jonathan A C Sterne, the Longitudinal Health and Wellbeing COVID-19 National Core Study

**Author notes:** **CORRESPONDING AUTHOR** Jonathan Sterne. authors contributed equally (Venexia M Walker, Praveetha Patalay, Jose Ignacio Cuitun Coronado). authors contributed equally (Nishi Chaturvedi, John Macleod, Ann John, Jonathan A C Sterne).

## Abstract

**Background:** COVID-19 is associated with subsequent mental illness in both hospital- and population-based studies. Evidence regarding effects of COVID-19 vaccination on mental health consequences of COVID-19 is limited.

**Methods:** With the approval of NHS England, we used linked electronic health records (OpenSAFELY-TPP) to conduct analyses in a ‘pre-vaccination’ cohort (17,619,987 people) followed during the wild-type/Alpha variant eras (January 2020-June 2021), and ‘vaccinated’ and ‘unvaccinated’ cohorts (13,716,225 and 3,130,581 people respectively) during the Delta variant era (June-December 2021). We estimated adjusted hazard ratios (aHRs) comparing the incidence of mental illness after diagnosis of COVID-19 with the incidence before or without COVID-19.

**Outcomes:** We considered eight outcomes: depression, serious mental illness, general anxiety, post-traumatic stress disorder, eating disorders, addiction, self-harm, and suicide. Incidence of most outcomes was elevated during weeks 1-4 after COVID-19 diagnosis, compared with before or without COVID-19, in each cohort. Vaccination mitigated the adverse effects of COVID-19 on mental health: aHRs (95% CIs) for depression and for serious mental illness during weeks 1-4 after COVID-19 were 1.93 (1.88-1.98) and 1.42 (1.24-1.61) respectively in the pre-vaccination cohort and 1.79 (1.68-1.91) and 2.21 (1.99-2.45) respectively in the unvaccinated cohort, compared with 1.16 (1.12-1.20) and 0.91 (0.84-0.98) respectively in the vaccinated cohort. Elevation in incidence was higher, and persisted for longer, after hospitalised than non-hospitalised COVID-19.

**Interpretation:** Incidence of mental illness is elevated for up to a year following severe COVID-19 in unvaccinated people. Vaccination mitigates the adverse effect of COVID-19 on mental health.

**Funding:** Medical Research Council (MC_PC_20059) and NIHR (COV-LT-0009).

## INTRODUCTION

SARS-CoV-2 infection, and consequent COVID-19, are associated with subsequent mental illness in both hospital and population-based studies^1,2^; for both common mental health difficulties, such as anxiety and depressive symptoms^3^, and serious mental illness, including psychotic disorders^4^. Potential mechanisms for these adverse effects of COVID-19 include physiological pathways, such as inflammation and microvascular changes, and psychosocial effects, such as anxiety about the consequences of COVID-19 including long COVID. Previous studies identified associations of COVID-19 with mental illness in both hospitalised patients^5,6^ and the general population^1,7,8^. Differentiating between hospitalised and non-hospitalised groups can provide insights into the role of COVID-19 severity.^9^

Rapid rollout of COVID-19 vaccination within a year of the start of the pandemic was a crucial component of the public health response. Although the impacts of vaccination in preventing and reducing the severity of COVID-19 are well-established^10,11^, there is limited evidence regarding the implications of vaccination for other adverse health consequences of COVID-19, including mental illness. Several studies reported short-term improvements in population mental health following vaccination rollout.^12,13^ However, we did not identify any studies investigating differences in mental illness outcomes following COVID-19 by vaccination status.

Rates of SARS-CoV-2 infection, vaccination, and disease severity were patterned by socio-demographic and health factors including age, sex, ethnicity, income and prior mental illness.^14–18^ Mental health consequences of COVID-19 may also vary between subgroups. For example, older people are at higher risk of more severe COVID-19, and hence may have experienced higher levels of psychological distress.

Using linked primary and secondary care data from over 17 million people, we examined associations of COVID-19 with subsequent mental illness in the pre-vaccination period of the pandemic and for unvaccinated and vaccinated people after vaccination became available. We compared rates of common and severe mental illness after a diagnosis of COVID-19 with rates before or without COVID-19. We also investigated variation in these associations between subgroups defined by COVID-19 severity, age, sex, ethnicity, prior mental illness, and prior SARS-COV-2 infection. Follow-up of those diagnosed with COVID-19 during the first year of the pandemic was for up to two years post-diagnosis.

## METHODS

### Study design and data sources

Our study used OpenSAFELY-TPP, which provides secure, privacy-protecting access to linked data from 24 million people registered with English general practices (GPs) using TPP SystmOne software. These data include primary care data linked via pseudonymised NHS number to Secondary Uses Service (SUS) secondary care data, Office of National Statistics (ONS) Death Registry, Second Generation Surveillance System (SGSS) COVID-19 testing data and the Index of Multiple Deprivation (IMD). COVID-19 vaccination records (National Immunisation Management System) are available within TPP primary care data.

Outcomes were defined using the earliest of: a relevant SNOMED CT code indicating a diagnosis in primary care; start of a secondary care episode with an ICD-10 code indicating a confirmed diagnosis in any position; or death with an ICD-10 code indicating the diagnosis as either a primary or underlying cause. From the range of mental illnesses examined, we present main findings for depression and serious mental illness (composite of schizophrenia, schizo-affective disorder, bipolar disorder, and psychotic depression). We also examined general anxiety disorders, post-traumatic stress disorder (PTSD), eating disorders, addiction, self-harm and suicide, which are presented in supplementary material. Code lists are available online: https://github.com/opensafely/post-covid-mentalhealth/tree/main/codelists.

The date of COVID-19 was defined as the first of: confirmed COVID-19 diagnosis recorded in primary care; positive SARS-COV-2 PCR or antigen test recorded in SGSS; start of an episode with a confirmed diagnosis in any position in SUS; or death with SARS-COV-2 infection listed in any position on the ONS death registry.

People with a hospital admission record in SUS that included a confirmed diagnosis in the primary position within 28 days of first COVID-19 were defined as having had ‘hospitalised COVID-19’. All other COVID-19 diagnoses were defined as ‘non-hospitalised’. Covariates identified as potential confounders included age, sex, ethnicity, area socioeconomic deprivation, smoking status, care home residence, health care work, number of GP-patient interactions in 2019, and binary indicators for history of comorbidities (**Supplementary Table 1**).

### Study population

Three cohorts were defined (**Supplementary Table 2; Supplementary Figure 1**). The ‘pre-vaccination’ cohort was followed from January 1^st^ 2020 (baseline) until the earliest of December 14^th^ 2021^19^, outcome event date and date of death. Exposure was defined as recorded COVID-19 between baseline and the earliest date of eligibility for COVID-19 vaccination, date of first vaccination and June 18^th^ 2021 (when all adults became eligible for vaccination). Follow-up in the ‘vaccinated’ cohort started at the later of baseline and two weeks after a second COVID-19 vaccination and ended at the earliest of December 14^th^ 2021, outcome event date and date of death. The ‘unvaccinated’ cohort had not received a COVID-19 vaccine by 12 weeks after they became eligible for vaccination. Follow-up started at the later of baseline and 12 weeks after vaccination eligibility and ended at the earliest of December 14^th^ 2021, outcome event date, date of death and date of first vaccination.

People eligible for each cohort were registered with an English GP for at least six months before baseline and were alive with a known age between 18 and 110 years, sex, deprivation, and region at baseline. People were excluded from each cohort if they had any record of SARS-CoV-2 infection before baseline. In the vaccinated cohort, people were excluded if they received a vaccination before the start of the vaccine rollout on December 8^th^ 2020 (indicating an error or participation in a randomized trial); their second dose was dated before their first dose vaccination (contradictory vaccine record); their second dose was less than three weeks after their first dose; or they received mixed vaccine brands before this was permitted on May 7^th^ 2021. In the unvaccinated cohort, people were excluded if they had any record of a COVID-19 vaccination before June 1^st^ 2021. Vaccine eligibility was defined using the Joint Committee on Vaccination and Immunisation (JCVI) groupings. People who could not be assigned to a JCVI group were excluded.

### Statistical analyses

For each cohort, baseline characteristics were described, and numbers of outcome events, person-years of follow-up and incidence rates (per 100,000 person-years) before and after all, hospitalised and non-hospitalised COVID-19 were tabulated. Time to first event was analysed for each outcome. Cox models were fitted with calendar time scale using the cohort-specific baseline as the origin. Hazard ratios (HRs) for follow-up after, versus before or without COVID-19, were estimated, splitting follow-up into the day of COVID-19 diagnosis (‘day 0’), the remainder of 1-4 weeks, and 5-28 weeks after COVID-19 for all cohorts and additionally 29-52 and 53-102 weeks after COVID-19 for the pre-vaccination cohort. For computational efficiency, we used sampling for analyses containing >4,000,000 people: we included all people with the outcome event, all people with the exposure, and a 10% (for general anxiety, depression, serious mental illness) or 20% (for all other outcomes) random sample of non-case-non-exposed people. We used inverse probability weights to adjust for the sampling and derived confidence intervals using robust standard errors when sampling had occurred. For each outcome and cohort, we estimated: (i) age and sex adjusted; and (ii) maximally adjusted HRs including all covariates listed above. Restricted cubic splines were used to account for age unless otherwise specified. All models were stratified by region so that risk sets were constructed within region, hence accounting for between-region variation in the baseline hazard.

Subgroup analyses according to prior history of the outcome event, age group, sex, ethnicity, and COVID-19 history were conducted for depression and serious mental illness. We calculated absolute excess risk 28 weeks after COVID-19, including outcome events recorded on the day of COVID-19 diagnosis (‘day 0’) and weighted by the proportion of people in age and sex strata in the pre-vaccination cohort. Further details of the statistical methods are in the supplement.

Data management and analyses were conducted in Python version 3.8.10, R version 4.0.2 and Stata/MP version 16.1 according to a pre-specified protocol. The protocol, analysis code and code lists are available online: https://github.com/opensafely/post-covid-mentalhealth.

## RESULTS

The pre-vaccination cohort included 17,619,987 people of whom 975,429 received a COVID-19 diagnosis (**Table 1, Supplementary Figure 2**). The median (interquartile range (IQR)) age was 49 (34-64) years. The cohort was 50.2% female and 79.3%, 6.3%, and 2.1% were recorded as of White, South Asian, and Black ethnicities respectively. The vaccinated and unvaccinated cohorts included 13,716,225 and 3,130,581 people respectively, of whom 850,023 and 147,315 received a COVID-19 diagnosis. Differences in demographic characteristics between the vaccinated and unvaccinated cohorts reflect predictors of COVID-19 vaccine uptake.^20^ **Supplementary Table 3** summarises participants’ medical history by cohort.

**Table 1:** Patient characteristics in the pre-vaccination, vaccinated and unvaccinated cohorts.

| Characteristic |  | Pre-vaccination cohort |  | Vaccinated cohort |  | Unvaccinated cohort |  |
| --- | --- | --- | --- | --- | --- | --- | --- |
|  |  | N (%) | COVID-19 diagnoses | N (%) | COVID-19 diagnoses | N (%) | COVID-19 diagnoses |
| All |  | 17619987 | 975429 | 13716225 | 850023 | 3130581 | 147315 |
| Sex | Female | 8847327 (50.2%) | 524127 | 7140045 (52.1%) | 472455 | 1313967 (42%) | 77943 |
|  | Male | 8772657 (49.8%) | 451299 | 6576177 (47.9%) | 377571 | 1816611 (58%) | 69375 |
| Age, years | 18-29 | 2968245 (16.8%) | 230823 | 1695351 (12.4%) | 92115 | 948093 (30.3%) | 39417 |
|  | 30-39 | 3018537 (17.1%) | 197505 | 1914075 (14%) | 146811 | 939573 (30%) | 49947 |
|  | 40-49 | 2921427 (16.6%) | 185643 | 2162535 (15.8%) | 232005 | 593535 (19%) | 32889 |
|  | 50-59 | 3135621 (17.8%) | 180213 | 2663187 (19.4%) | 200301 | 361947 (11.6%) | 16545 |
|  | 60-69 | 2452011 (13.9%) | 90525 | 2270565 (16.6%) | 103599 | 175419 (5.6%) | 5535 |
|  | 70-79 | 1990863 (11.3%) | 49467 | 1940685 (14.1%) | 53013 | 76089 (2.4%) | 1899 |
|  | 80-89 | 940581 (5.3%) | 29667 | 889707 (6.5%) | 17853 | 28389 (0.9%) | 879 |
|  | 90+ | 192699 (1.1%) | 11583 | 180123 (1.3%) | 4323 | 7527 (0.2%) | 219 |
| Ethnicity | White | 13965363 (79%) | 737295 | 11361897 (83%) | 726897 | 1946799 (62%) | 112497 |
|  | S. Asian | 1109529 (6.3%) | 114231 | 769293 (5.6%) | 40809 | 311121 (9.9%) | 9381 |
|  | Black | 368835 (2.1%) | 25845 | 212187 (1.5%) | 9405 | 166593 (5.3%) | 7443 |
|  | Other | 371805 (2.1%) | 19137 | 228615 (1.7%) | 10377 | 180609 (5.8%) | 3951 |
|  | Mixed | 197745 (1.1%) | 13467 | 124425 (0.9%) | 7383 | 77583 (2.5%) | 3789 |
|  | Missing | 1606707 (9.1%) | 65451 | 1019811 (7.4%) | 55149 | 447873 (14.3%) | 10257 |
| IMD quintile (lower: more deprived) | 1 | 3395079 (19.3%) | 239709 | 2229885 (16.3%) | 133593 | 933093 (29.8%) | 45183 |
|  | 2 | 3478545 (19.7%) | 211725 | 2563539 (18.7%) | 156321 | 750447 (24%) | 34959 |
|  | 3 | 3801099 (21.6%) | 194859 | 3032523 (22.1%) | 183045 | 627477 (20%) | 29079 |
|  | 4 | 3606213 (20.5%) | 176199 | 2999331 (21.9%) | 188019 | 479739 (15.3%) | 22113 |
|  | 5 | 3339045 (19%) | 152931 | 2890947 (21.1%) | 189039 | 339819 (10.9%) | 15981 |
| Smoking | Never | 682269 (3.9%) | 38217 | 412623 (3%) | 15945 | 356319 (11.4%) | 8247 |
|  | Former | 8051205 (45.7%) | 485781 | 6381579 (46.5%) | 414003 | 1295349 (41.4%) | 58611 |
|  | Current | 5870427 (33.3%) | 318777 | 5000223 (36.5%) | 327639 | 625515 (20%) | 43167 |
|  | Missing | 3016083 (17.1%) | 132645 | 1921797 (14%) | 92433 | 853395 (27.3%) | 37287 |
| Region | East | 4088403 (23.2%) | 221181 | 3209673 (23.4%) | 180231 | 704565 (22.5%) | 33441 |
|  | E.Midlands | 3121905 (17.7%) | 185739 | 2431149 (17.7%) | 158091 | 513993 (16.4%) | 28311 |
|  | London | 1116735 (6.3%) | 64383 | 715353 (5.2%) | 37059 | 446763 (14.3%) | 10929 |
|  | North East | 852093 (4.8%) | 58887 | 652491 (4.8%) | 48537 | 132987 (4.2%) | 7185 |
|  | North West | 1568139 (8.9%) | 104913 | 1233741 (9%) | 85629 | 211239 (6.7%) | 11697 |
|  | South East | 1159293 (6.6%) | 47829 | 943521 (6.9%) | 55689 | 188109 (6%) | 8757 |
|  | South West | 2490369 (14.1%) | 76029 | 2136165 (15.6%) | 129291 | 313809 (10%) | 17235 |
|  | W.Midlands | 693339 (3.9%) | 52617 | 481725 (3.5%) | 29205 | 172029 (5.5%) | 7941 |
|  | Yorkshire | 2529699 (14.4%) | 163839 | 1912395 (13.9%) | 126285 | 447087 (14.3%) | 21819 |
| Care home resident |  | 82575 (0.5%) | 14391 | 56451 (0.4%) | 2775 | 2889 (0.1%) | 105 |
| Healthcare worker |  | 552441 (3.1%) | 63351 | 480825 (3.5%) | 38709 | 20157 (0.6%) | 1947 |

IMD: Index of Multiple Deprivation

In each cohort, the incidence of mental illness was higher after diagnosis of COVID-19 than before or without COVID-19 (**Table 2**). The highest incidence rates were after hospitalised COVID-19. Depression was the most common outcome with a total of 1,278,363, 343,371, and 56,403 diagnoses in the pre-vaccination, vaccinated and unvaccinated cohorts respectively. There were 379,275, 84,981, and 18,039 diagnoses of serious mental illness in the pre-vaccination, vaccinated and unvaccinated cohorts respectively.

**Table 2:** Mental illness events following diagnosis of COVID-19 in the pre-vaccination, vaccinated and unvaccinated cohorts, overall and by COVID-19 severity.

| Outcome | COVID-19 severity | Pre-vaccination cohort<br>N=17,619,987 |  | Vaccinated cohort<br>N=13,716,225 |  | Unvaccinated cohort<br>N=3,130,581 |  |
| --- | --- | --- | --- | --- | --- | --- | --- |
|  |  | Event/person-years | Incidence rate | Event/person-years | Incidence rate | Event/person-years | Incidence rate |
| Depression | No COVID-19 | 1229103/31815936 | 3863 | 332523/6177768 | 5383 | 53547/1178058 | 4545 |
|  | Hospitalised COVID-19 | 6123/40514 | 15113 | 1359/2379 | 57128 | 831/1614 | 51475 |
|  | Non-hospitalised COVID-19 | 43137/875165 | 4929 | 9489/153403 | 6186 | 2025/24665 | 8210 |
| Serious mental illness | No COVID-19 | 364809/32692824 | 1116 | 82479/6242112 | 1321 | 17325/1185912 | 1461 |
|  | Hospitalised COVID-19 | 1581/46044 | 3434 | 207/2651 | 7809 | 165/1771 | 9319 |
|  | Non-hospitalised COVID-19 | 12885/921950 | 1398 | 2295/156141 | 1470 | 549/25091 | 2188 |
| General anxiety disorders | No COVID-19 | 898383/32181914 | 2792 | 221499/6207309 | 3568 | 39285/1181444 | 3325 |
|  | Hospitalised COVID-19 | 4695/42625 | 11015 | 1017/2481 | 40994 | 873/1615 | 54045 |
|  | Non-hospitalised COVID-19 | 34569/891502 | 3878 | 6795/154561 | 4396 | 1689/24774 | 6818 |
| Post-traumatic stress disorder | No COVID-19 | 35325/33027918 | 107 | 8061/6260421 | 129 | 2733/1189246 | 230 |
|  | Hospitalised COVID-19 | 303/47761 | 634 | 57/2695 | 2115 | 57/1798 | 3170 |
|  | Non-hospitalised COVID-19 | 1281/939590 | 136 | 225/156887 | 143 | 93/25218 | 369 |
| Eating disorders | No COVID-19 | 18273/33044280 | 55 | 4539/6261356 | 72 | 903/1189653 | 76 |
|  | Hospitalised COVID-19 | 63/48018 | 131 | Sep-07 | 333 | 15/1808 | 830 |
|  | Non-hospitalised COVID-19 | 765/940251 | 81 | 129/156909 | 82 | 33/25239 | 131 |
| Addiction | No COVID-19 | 36615/33023855 | 111 | 5481/6261035 | 88 | 4131/1188788 | 347 |
|  | Hospitalised COVID-19 | 165/47887 | 345 | 21/2704 | 777 | 51/1799 | 2835 |
|  | Non-hospitalised COVID-19 | 879/939847 | 94 | 105/156925 | 67 | 87/25220 | 345 |
| Self-harm | No COVID-19 | 85761/32977385 | 260 | 15027/6258886 | 240 | 4977/1188790 | 419 |
|  | Hospitalised COVID-19 | 237/47754 | 496 | 27/2703 | 999 | 15/1808 | 830 |
|  | Non-hospitalised COVID-19 | 2955/936599 | 316 | 507/156793 | 323 | 141/25205 | 559 |
| Suicide | No COVID-19 | 39951/33023812 | 121 | 6135/6260977 | 98 | 1935/1189437 | 163 |
|  | Hospitalised COVID-19 | 159/47888 | 332 | 15/2706 | 554 | 9/1810 | 497 |
|  | Non-hospitalised COVID-19 | 1359/939065 | 145 | 219/156893 | 140 | 57/25232 | 226 |

### Comparisons of event rates after diagnosis of COVID-19 versus before or without COVID-19

Maximally adjusted HRs (aHRs) comparing the incidence of each outcome after diagnosis of COVID-19 with the incidence before or without COVID-19 did not differ substantially from the age- and sex-adjusted HRs in all cohorts (**Supplementary Figure 3**). The incidence of all outcomes was extremely high on day zero (**Table 3**). The incidence of most outcomes was elevated during the remainder of 1-4 weeks after COVID-19, compared with before or without COVID-19, in each cohort.

**Table 3:** Maximally adjusted hazard ratios and 95% CIs for mental illness events following diagnosis of COVID-19 in the pre-vaccination, vaccinated and unvaccinated cohorts.

| Outcome | Time since COVID-19 | Pre-vaccination cohort | Vaccinated cohort | Unvaccinated cohort |
| --- | --- | --- | --- | --- |
| Depression | Day 0 | 32.9 (31.8-34.0) | 8.52 (8.05-9.03) | 19.1 (17.5-20.8) |
|  | 1-4 weeks | 1.93 (1.88-1.98) | 1.16 (1.12-1.20) | 1.79 (1.68-1.91) |
|  | 5-28 weeks | 1.25 (1.23-1.26) | 1.11 (1.08-1.14) | 1.28 (1.22-1.36) |
|  | 29-52 weeks | 1.16 (1.14-1.18) | - | - |
|  | 53-102 weeks | 1.17 (1.13-1.20) | - | - |
| Serious mental illness | Day 0 | 29.4 (27.6-31.3) | 7.61 (6.74-8.59) | 19.1 (16.2-22.5) |
|  | 1-4 weeks | 1.48 (1.40-1.56) | 0.91 (0.84-0.98) | 1.42 (1.24-1.61) |
|  | 5-28 weeks | 1.20 (1.17-1.23) | 1.07 (1.01-1.12) | 1.14 (1.02-1.27) |
|  | 29-52 weeks | 1.15 (1.12-1.18) | - | - |
|  | 53-102 weeks | 1.13 (1.06-1.19) | - | - |
| General anxiety | Day 0 | 33.7 (32.4-34.9) | 8.55 (7.98-9.17) | 25.3 (23.3-27.5) |
|  | 1-4 weeks | 2.19 (2.13-2.26) | 1.26 (1.21-1.31) | 2.29 (2.15-2.44) |
|  | 5-28 weeks | 1.26 (1.24-1.28) | 1.12 (1.09-1.16) | 1.36 (1.28-1.45) |
|  | 29-52 weeks | 1.14 (1.12-1.16) | - | - |
|  | 53-102 weeks | 1.15 (1.11-1.19) | - | - |
| Post-traumatic stress disorder | Day 0 | 43.3 (36.8-51.0) | 12.3 (9.13-16.4) | 30.0 (22.1-40.5) |
|  | 1-4 weeks | 1.84 (1.56-2.16) | 1.01 (0.81-1.26) | 1.42 (1.06-1.92) |
|  | 5-28 weeks | 1.13 (1.04-1.23) | 1.01 (0.86-1.18) | 1.05 (0.81-1.36) |
|  | 29-52 weeks | 1.06 (0.97-1.16) | - | - |
|  | 53-102 weeks | 1.17 (0.99-1.38) | - | - |
| Eating disorders | Day 0 | 29.2 (22.7-37.6) | 5.59 (3.26-9.59) | 20.9 (11.8-37.0) |
|  | 1-4 weeks | 1.96 (1.61-2.39) | 0.88 (0.64-1.21) | 1.56 (0.97-2.52) |
|  | 5-28 weeks | 1.26 (1.13-1.41) | 0.91 (0.71-1.16) | 1.14 (0.73-1.77) |
|  | 29-52 weeks | 1.09 (0.96-1.24) | - | - |
|  | 53-102 weeks | 1.24 (0.98-1.57) | - | - |
| Addiction | Day 0 | 66.2 (56.9-77.1) | 17.0 (12.0-24.1) | 44.8 (34.9-57.5) |
|  | 1-4 weeks | 1.45 (1.19-1.78) | 0.98 (0.72-1.33) | 1.04 (0.73-1.47) |
|  | 5-28 weeks | 0.95 (0.86-1.05) | 0.69 (0.53-0.91) | 0.77 (0.57-1.05) |
|  | 29-52 weeks | 1.00 (0.89-1.12) | - | - |
|  | 53-102 weeks | 1.00 (0.79-1.26) | - | - |
| Self-harm | Day 0 | 22.3 (19.2-26.0) | 10.9 (8.61-13.9) | 12.1 (8.28-17.6) |
|  | 1-4 weeks | 1.21 (1.06-1.37) | 1.02 (0.86-1.20) | 1.09 (0.83-1.44) |
|  | 5-28 weeks | 1.15 (1.09-1.22) | 1.17 (1.04-1.31) | 0.92 (0.73-1.15) |
|  | 29-52 weeks | 1.17 (1.10-1.25) | - | - |
|  | 53-102 weeks | 1.14 (1.01-1.29) | - | - |
| Suicide | Day 0 | 850 (742-975) | † | † |
|  | 1-4 weeks | 4.18 (2.94-5.94) | † | † |
|  | 5-28 weeks | 0.85 (0.60-1.19) | † | † |
|  | 29-52 weeks | 0.83 (0.57-1.20) | - | - |
|  | 53-102 weeks | 0.86 (0.45-1.65) | - | - |

† Insufficient events for estimation

#### Depression

The incidence of depression was elevated during weeks 1-4 after COVID-19, compared with before or without COVID-19, in the pre-vaccination and unvaccinated cohorts (1.93 (95% CI 1.88-1.98) and 1.79 (1.68-1.91) respectively) and, to a lesser extent, in the vaccinated cohort (1.16 (1.12-1.20)) (**Figure 1, Table 3**). The incidence of depression remained elevated during weeks 5-28 in the vaccinated and unvaccinated cohorts (aHRs 1.11 (1.08-1.14) and 1.28 (1.22-1.36) respectively) and up to weeks 53-102 in the pre-vaccination cohort (aHR 1.17 (1.13-1.20)). aHRs during weeks 1-4 were considerably higher after hospitalised COVID-19 (pre-vaccination: 16.5 (15.8-17.3); vaccinated: 13.0 (12.0-14.0); unvaccinated: 15.6 (14.1-17.2)) than after non-hospitalised COVID-19 (pre-vaccination: 1.22 (1.17-1.26); vaccinated: 0.92 (0.88-0.95); unvaccinated: 1.11 (1.02-1.20)) (**Figure 1, Supplementary Tables 4-5**). In the pre-vaccination cohort, aHRs remained higher after hospitalised than non-hospitalised COVID-19 throughout follow-up.

**Figure 1:**
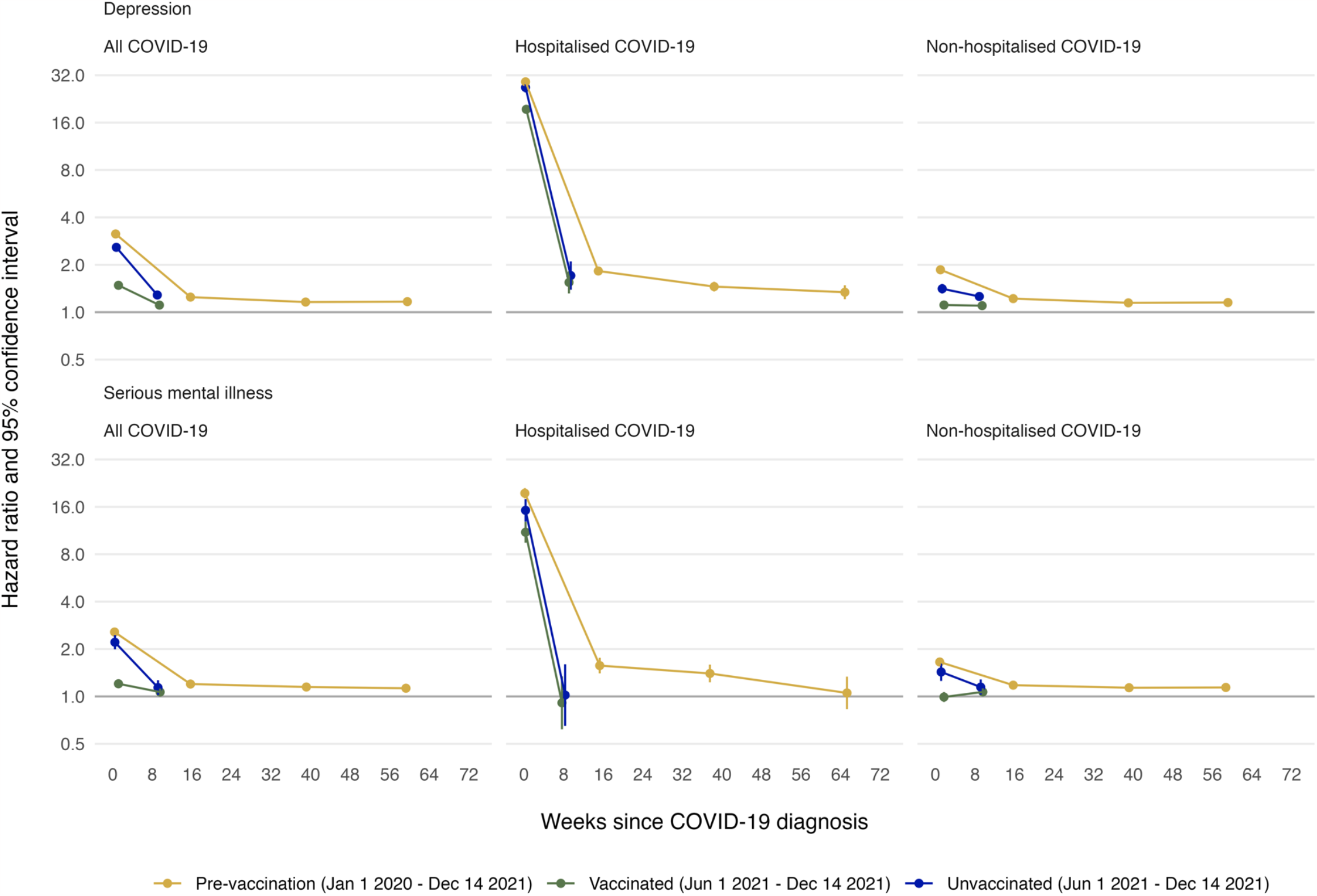
Maximally adjusted hazard ratios and 95% CIs for depression and serious mental illness following diagnosis of COVID-19 in the pre-vaccination, vaccinated and unvaccinated cohorts, overall and by COVID-19 severity. Events on the day of COVID-19 diagnosis (day 0) were excluded.

#### Serious mental illness

The incidence of serious mental illness was elevated during weeks 1-4 after COVID-19, compared with before or without COVID-19, in the pre-vaccinated and unvaccinated cohorts (1.48 (95% CI 1.40-1.56) and 1.42 (1.24-1.61) respectively) (**Figure 1, Table 3**). However, the incidence of serious mental illness was lower during weeks 1-4 in the vaccinated cohort (0.91 (0.84-0.98)). Incidence remained slightly elevated during weeks 5-28 in the vaccinated and unvaccinated cohorts (1.07 (1.01-1.1) and 1.14 (1.02-1.27)) and up to weeks 53-102 in the pre-vaccination cohort (1.13 (1.06-1.19)). The incidence of serious mental illness during weeks 1-4 was considerably higher after hospitalised COVID-19 (pre-vaccination: 9.65 (8.71-10.7); vaccinated: 6.38 (5.21-7.80); unvaccinated: 8.76 (7.02-10.9)) than after non-hospitalised COVID-19: (pre-vaccination: 1.04 (0.97-1.11); vaccinated: 0.79 (0.73-0.86); unvaccinated: 0.98 (0.83-1.15)) (**Figure 1, Supplementary Tables 4-5**).

#### Subgroup analyses

aHRs for depression were highest during weeks 1-4 after COVID-19, versus before or without COVID-19, for people with prior history of the condition that was recorded more than six months ago (**Figure 2, Supplementary Tables 6-8**). For example, in the pre-vaccination cohort, aHRs were 2.16 (2.08-2.23) for prior history more than six months ago, 1.66 (1.52-1.80) for prior history within six months, and 1.58 (1.50-1.67) for no prior history. aHRs for serious mental illness were also highest during weeks 1-4 after COVID-19 for people with prior history of the condition that was recorded more than six months ago in the unvaccinated cohort (1.56 (1.30-1.88)). However, aHRs for serious mental illness were highest during weeks 1-4 after COVID-19 for people with prior history within six months for the pre-vaccination (1.88 (1.57-2.26)) and vaccinated cohorts (1.07 (0.87-1.33)). Beyond 5 weeks, incidence attenuated for those with prior history both within six months and more than six months ago. For the vaccinated cohort, incidence of serious mental illness during weeks 5-28 after COVID-19 was consistent with before or without COVID-19 (prior history more than six months ago: 1.00 (0.93-1.08); prior history within six months: 1.07 (0.87-1.33)).

**Figure 2:**
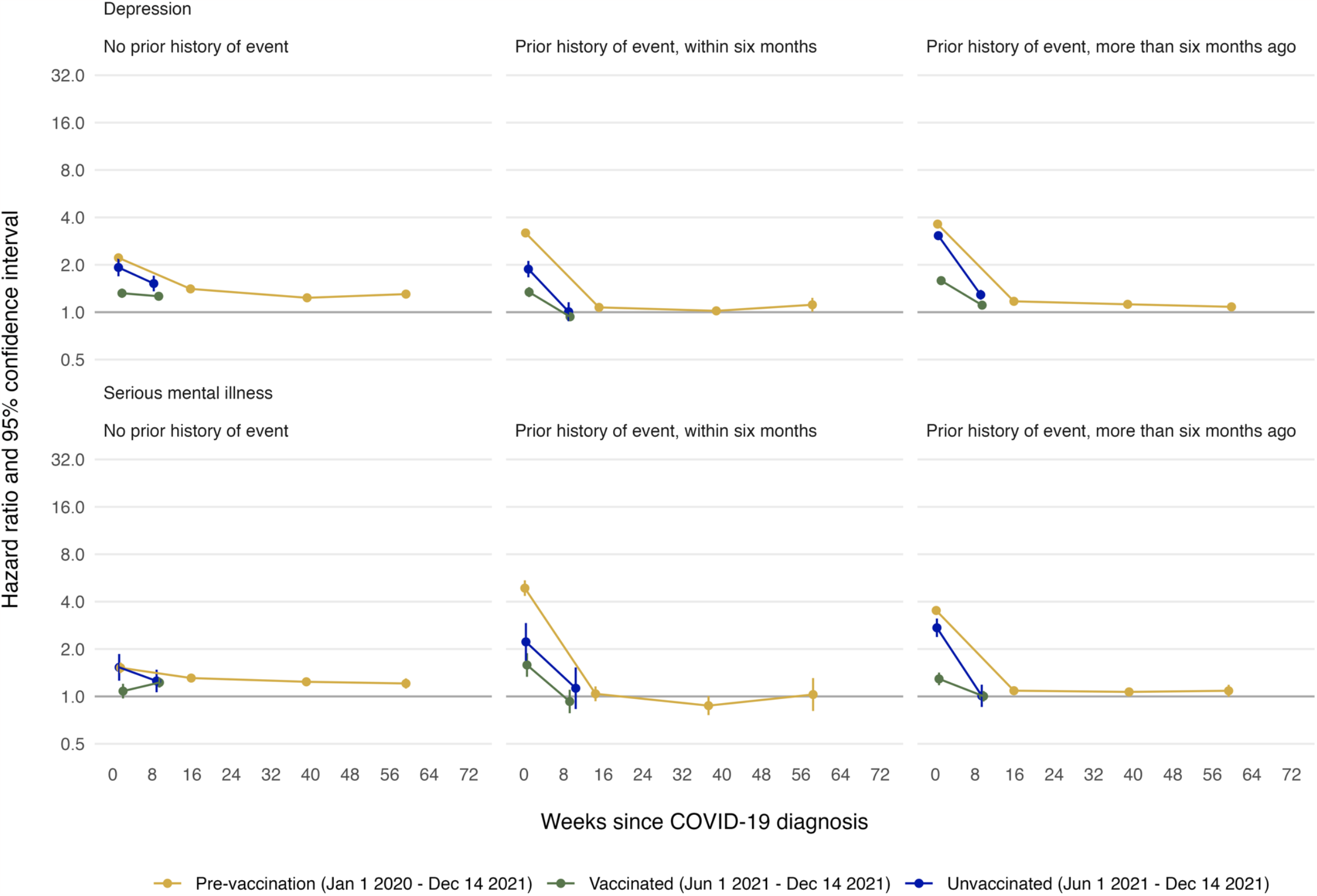
Maximally adjusted hazard ratios and 95% CIs for depression and serious mental illness following diagnosis of COVID-19 in the pre-vaccination, vaccinated and unvaccinated cohorts, by prior history of the outcome. Events on the day of COVID-19 diagnosis (day 0) were excluded.

In the vaccinated cohort, aHRs for depression and serious mental illness after COVID-19, versus before or without COVID-19 were similar in people with and without a with prior history of COVID-19 (**Supplementary Figure 4, Supplementary Table 9**). aHRs for depression during weeks 1-4 and 5-12 were greater in the 60-79 and 80-110 age groups than the 18-39 and 40-59 age groups: those serious mental illness were greater in the 60-79 and 80-110 age groups than the 18-39 and 40-59 age groups across all time periods (**Supplementary Figure 5, Supplementary Tables 10-13**). aHRs for depression and serious mental illness were marginally higher for men than women during weeks 1-4 after COVID-19 (**Supplementary Figure 6, Supplementary Tables 14-15**). aHRs for depression after COVID-19, versus before or without COVID-19, were generally comparable between ethnic groups (**Supplementary Figure 7, Supplementary Tables 16-20**), except that in the vaccinated cohort aHRs for depression were higher for the Black ethnic group than other ethnic groups. aHRs for serious mental illness after COVID-19 were broadly comparable by ethnic group for the pre-vaccination cohort. aHRs for serious mental illness could only be estimated for the White and South Asian ethnic groups in the vaccinated cohort, and for the White ethnic group in the unvaccinated cohort, due to low event counts.

#### Other mental illnesses

aHRs for other mental illnesses were broadly similar to those for depression and serious mental illness, both overall (**Figure 3, Table 3**) and for hospitalised and non-hospitalised COVID-19 (**Supplementary Tables 4-5**). An exception was that aHRs for post-traumatic stress disorder after hospitalised COVID-19, versus before or without COVID-19, were higher during weeks 1-4 in the vaccinated cohort than the other two cohorts (pre-vaccination: 19.9 (15.6-25.5); vaccinated: 26.3 (19.3-35.8); unvaccinated: 14.0 (8.40-23.4)). This pattern was not present for non-hospitalised COVID-19 or overall.

**Figure 3:**
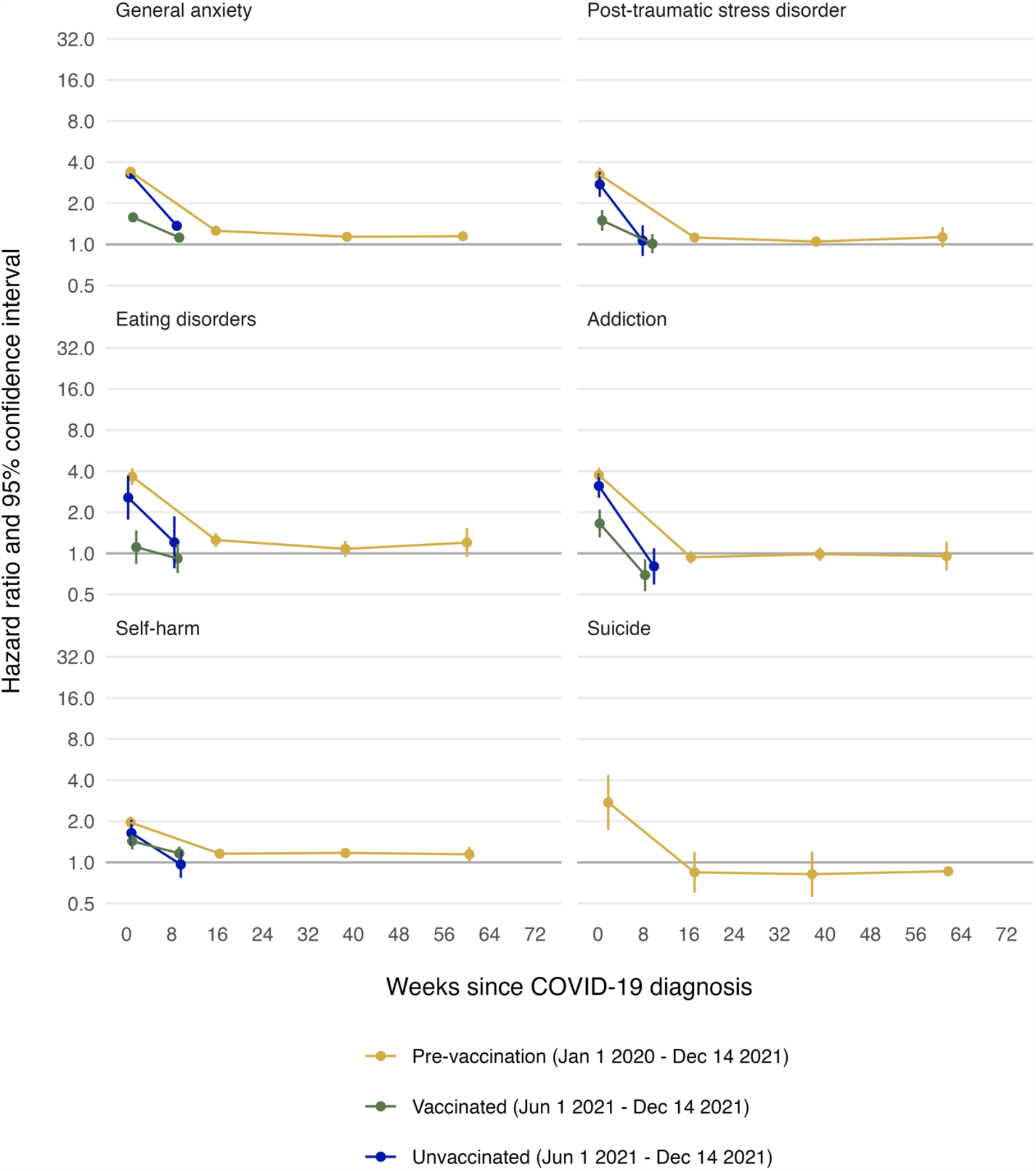
Maximally adjusted hazard ratios and 95% confidence intervals for other mental illness events following diagnosis of COVID-19 in the pre-vaccination, vaccinated and unvaccinated cohorts. Events on the day of COVID-19 diagnosis (day 0) were excluded.

### Absolute excess risk

Estimated excess risks of depression 28 weeks after COVID-19, standardised to the age and sex distribution of the pre-vaccination cohort, were 1020, 449, and 1009 per 100,000 people in the pre-vaccination, vaccinated and unvaccinated cohorts respectively (**Figure 4, Supplementary Table 21**). Up to 32% of the estimated excess events occurred on the day of COVID-19 diagnosis (‘day 0’). Estimated excess risks of serious mental illness 28 weeks post-COVID-19, standardised to the age and sex distribution of the pre-vaccination cohort, were 227, 60, and 199 per 100,000 people in the pre-vaccination, vaccinated and unvaccinated cohorts respectively. Up to 43% of the estimated excess events occured on the day of COVID-19 diagnosis (‘day 0’).

**Figure 4:**
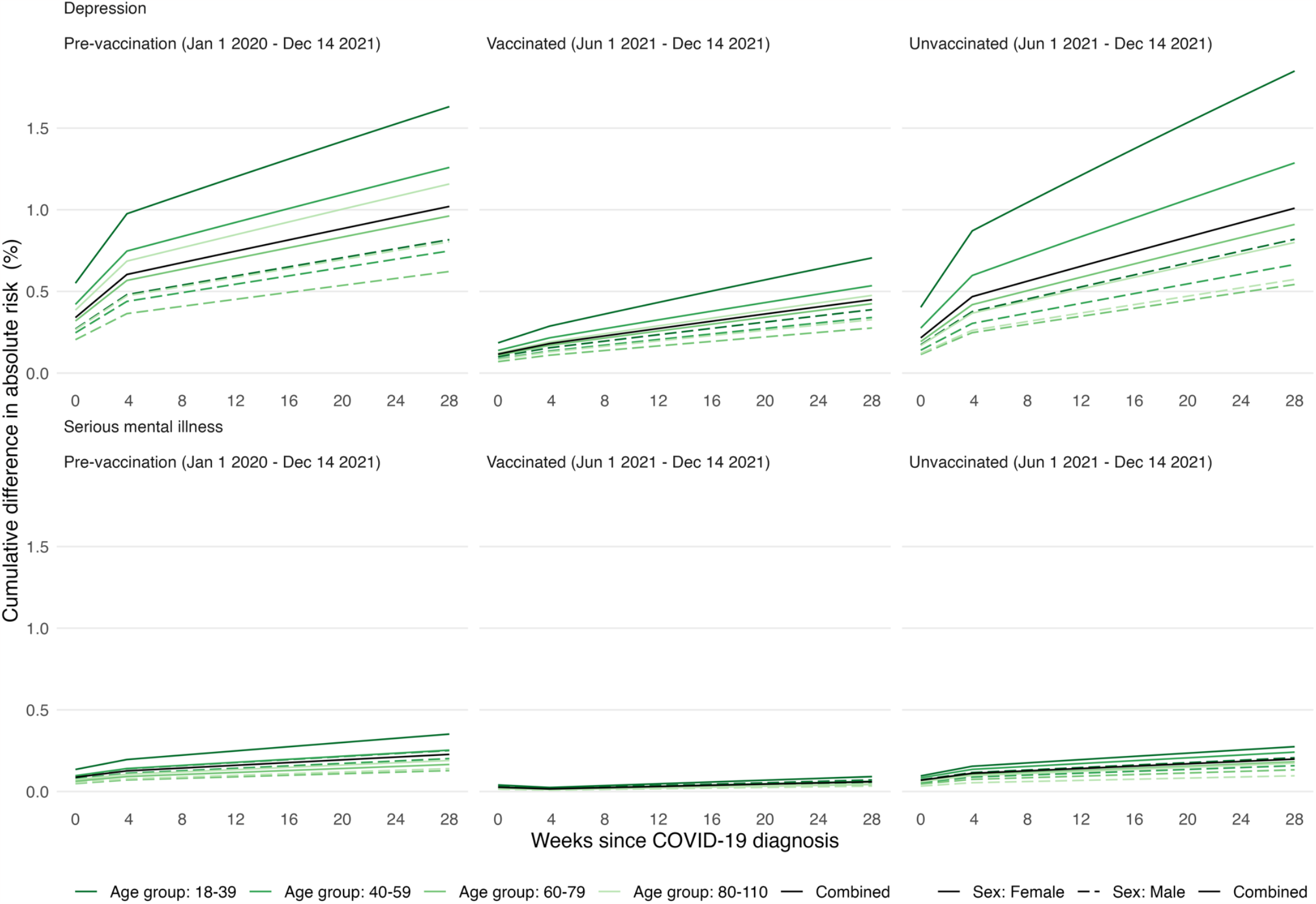
Absolute excess risk up to 28 weeks for depression and serious mental illness following diagnosis of COVID-19 in the pre-vaccination, vaccinated and unvaccinated cohorts.

## DISCUSSION

In a study of more than 17 million people followed for up to two years of the pandemic, rates of most mental illnesses were markedly elevated during the first four weeks after diagnosis of COVID-19. This elevation was less marked in people who were vaccinated before diagnosis of COVID-19. In people diagnosed with COVID-19 before vaccination was available, incidence of mental illness remained elevated more than four weeks after diagnosis, particularly in people who were hospitalised with COVID-19. In subgroup analyses according to prior history of depression and serious mental illness, associations 1-4 weeks after COVID-19 were greater in those with than without prior history of each outcome, but more than 4 weeks after COVID-19 associations were greater in people with no prior history of the outcome. Subgroup analyses also suggested stronger associations in older age groups and in men. The effect of COVID-19 on mental illness did not differ markedly between ethnic groups.

The attenuation of adverse effects of COVID-19 on mental illness in those who were vaccinated, compared to those unvaccinated, may be explained by reduced disease severity due to vaccination.^21^ This might be due to lower levels of systemic inflammation, as well as psychological benefits of vaccination such as reduced worry about the consequences of COVID-19, increased social engagement, and resuming previous activities.^22^ A previous cross-cohort study found that associations varied by COVID-19 severity, with poorer mental illness outcomes only found among those who were bedridden with COVID-19, as a marker of more severe illness.^3^ Those hospitalised due to COVID-19, especially people in intensive care and requiring ventilator support, may have been at greater risk of developing post-traumatic stress disorder^23^; though this could not be examined in the present study.

Elevation in rates of mental illness outcomes declined with increasing time since diagnosis of COVID-19, although for those diagnosed in in the pre-vaccination era incidence remained elevated up to a year after hospitalised COVID-19. Previous findings regarding long-term effects have been mixed, with a review reporting no clear long-term associations between COVID-19 and mental illness^24^, whereas a recent multi-cohort study found an association between COVID-19 and mental illness with little evidence of attenuation over time^1^. Persisting effects of COVID-19 on mental illness could partly reflect ongoing impacts of long COVID^25,26^.

In line with previous research^1^, we found stronger associations between COVID-19 and mental illness among older age groups. This is likely to reflect the increased risk of severe COVID-19 among older people and resulting increased anxiety about the consequences of infection. The association between COVID-19 and mental illness was slightly stronger among men, who have been found to be at greater risk of severe mental illness outcomes than women^27^. These patterns contrast with the wider impacts of the pandemic on mental health. For instance, overall mental health impacts of the pandemic were found to be greatest in adults aged 25-44 years, women, and those with higher degrees^28^; indicating that the mechanisms linking COVID-19 disease and mental health may differ from those underpinning wider effects of the pandemic.

Our findings highlight the wider public health benefits of the vaccination programme. Prior mental illness may influence vaccine uptake, which highlights the importance of actively encouraging vaccine uptake in people with mental health difficulties.^20,29,30^ Our analyses suggested that the adverse effects of COVID-19 on mental illness were greater during the first year of the pandemic, prior to the availability of vaccination. This may reflect greater uncertainty and public concern around consequences of COVID-19 and treatment effectiveness at the beginning of the pandemic.

Strengths of this study include the very large sample size, availability of detailed linked electronic health record data, relatively long duration of follow-up, and the opportunity to examine the role of vaccination in the relationship between COVID-19 and mental illness. We also note several limitations. First, those who were unvaccinated may have been less likely to contact health services and to test for SARS-CoV-2, which might have led to underestimated effects in unvaccinated people not hospitalised with COVID-19. Those with recorded COVID-19, particularly those who were hospitalised, may have been more likely to have their mental illness recorded due to greater contact with health services. This may have underpinned the particularly high HRs observed initially following diagnosis, especially in those hospitalised, and the rapid fall thereafter as service contact is likely to be highest in the early post-diagnosis period. However, this is unlikely to fully explain the adverse effect of COVID-19 on mental illness, given the persistent elevation of incidence of mental illness following hospitalised COVID-19 and the variation in associations with different mental illnesses. Also, those with prior recorded mental health diagnoses may not have had additional diagnostic codes added to their record at every visit, even if their mental health had deteriorated due to COVID-19. Additionally, data on mental health in primary and secondary care is generally incomplete, as it does not include access to mental health services data or NHS Talking Therapies (formerly ‘Improving Access to Psychological Therapies (IAPT)’), which patients can self-refer to. A further limitation is that we could only assess COVID-19 severity according to whether patients were hospitalised and did not consider the potential role of repeated infections. We cannot exclude the possibility of unmeasured confounding, although we controlled for a wide range of demographic characteristics and prior morbidities. A previous study found that the mental health impacts of COVID-19 were less apparent when using a negative control group, suggesting that observed associations may have been, at least in part, due to residual confounding^2^. Finally, other viruses may have consequences for mental illness. Our findings may therefore reflect a phenomenon that occurs after many viruses rather than being specific to SARS-CoV-2.

Our findings add to a growing body of evidence highlighting the increased risk of mental illness following COVID-19 diagnosis, with stronger associations found in relation to non-vaccination and more severe COVID-19 disease, and longer-term associations relating mainly to new-onset mental illness. This has important implications for public health and mental health service provision, as serious mental illnesses in particular are associated with more intensive healthcare needs and longer-term health and other adverse effects. Our results highlight the importance of accessing vaccination among those with mental illness, who may be at higher risk of both SARS-CoV-2 infection and adverse health outcomes following COVID-19. They also emphasise the widespread public health benefits of COVID-19 vaccination in the general population.

## ACKNOWLEDGEMENTS

We are very grateful for all the support received from the TPP Technical Operations team throughout this work, and for generous assistance from the information governance and database teams at NHS England and the NHS England Transformation Directorate. We thank the CONVALESCENCE Study Long COVID PPIE group for their input and for sharing their experiences and expertise throughout the duration of the project.

## DATA AVAILABILITY STATEMENT

All data were linked, stored and analysed securely within the OpenSAFELY platform: https://www.opensafely.org/. Data include pseudonymised data such as coded diagnoses, medications and physiological parameters. No free text data are included. All code and code lists are shared openly for review and re-use under an MIT open license (https://github.com/opensafely/post-covid-mentalhealth). Detailed pseudonymised patient data is potentially re-identifiable and therefore not shared.

## ETHICAL APPROVAL AND INFORMATION GOVERNANCE

This study was approved by the Health Research Authority [REC reference 22/PR/0095] and by the University of Bristol’s Faculty of Health Sciences Ethics Committee [reference 117269]. Authors involved in data management/analysis successfully passed information governance training and obtained ONS safe researcher accreditation. NHS England is the data controller of OpenSAFELY-TPP. All outputs underwent disclosure checks and were approved by NHS England. Further details of OpenSAFELY information governance are provided in supplemental methods.

## AUTHOR CONTRIBUTIONS

VW, PP, RD, AWo, NC, JMac, AJ, JACS contributed to Conceptualization. VW, RD, JACS contributed to Methodology. VW, JICC, TP, AWa, LF, JMas, SD, AM, SB, BG contributed to Software. VW, JICC, RD, HF, JS, BM, ET, KT, GC, EH, YW, MAA, RK, JMac, AJ contributed to Validation. VW, JICC contributed to Formal analysis. VW, JICC contributed to Investigation. TP, AWa, LF, JMas, SD, AM, SB, BG contributed to Resources. VW, JICC, HF contributed to Data Curation. VW, PP, JACS contributed to Writing - Original Draft. All authors contributed to Writing - Review & Editing. VW, JICC contributed to Visualization. VW, RD, BG, AWo, NC, JMac, AJ, JACS contributed to Supervision. VW, PP, RD, BG, AWo, NC, JMac, AJ, JACS contributed to Project administration. NC, JACS contributed to Funding acquisition.

## REFERENCES

1 Thompson EJ, Stafford J, Moltrecht B, et al. Psychological distress, depression, anxiety, and life satisfaction following COVID-19 infection: evidence from 11 UK longitudinal population studies. The Lancet Psychiatry 2022; 9: 894–906.

2 Abel KM, Carr MJ, Ashcroft DM, et al. Association of SARS-CoV-2 Infection With Psychological Distress, Psychotropic Prescribing, Fatigue, and Sleep Problems Among UK Primary Care Patients. JAMA Netw Open 2021; 4: e2134803.

3 Magnúsdóttir I, Lovik A, Unnarsdóttir AB, et al. Acute COVID-19 severity and mental health morbidity trajectories in patient populations of six nations: an observational study. The Lancet Public Health 2022; 7: e406–16.

4 Taquet M, Geddes JR, Husain M, Luciano S, Harrison HarrisonJ. 6-month neurological and psychiatric outcomes in 236 379 survivors of COVID-19: a retrospective cohort study using electronic health records. The Lancet Psychiatry 2021; 8: 416–27.

5 Taquet M, Dercon Q, Luciano S, Geddes JR, Husain M, Harrison HarrisonJ. Incidence, co-occurrence, and evolution of long-COVID features: A 6-month retrospective cohort study of 273,618 survivors of COVID-19. PLoS Med 2021; 18: e1003773.

6 Oh TK, Park HY, Song I. Risk of psychological sequelae among coronavirus disease-2019 survivors: A nationwide cohort study in South Korea. Depression and Anxiety 2021; 38: 247–54.

7 Klaser K, Thompson EJ, Nguyen LH, et al. Anxiety and depression symptoms after COVID-19 infection: results from the COVID Symptom Study app. J Neurol Neurosurg Psychiatry 2021; 92: 1254–8.

8 Niedzwiedz CL, Benzeval M, Hainey K, Leyland AH, Katikireddi KatikireddiV. Psychological distress among people with probable COVID-19 infection: analysis of the UK Household Longitudinal Study. BJPsych open 2021; 7: e104.

9 Nersesjan V, Christensen RHB, Kondziella D, Benros BenrosE. COVID-19 and Risk for Mental Disorders Among Adults in Denmark. JAMA Psychiatry 2023; 80: 778.

10 Mascellino MT, Di Timoteo F, De Angelis M, Oliva A. Overview of the Main Anti-SARS-CoV-2 Vaccines: Mechanism of Action, Efficacy and Safety. IDR 2021; Volume 14: 3459–76.

11 Suthar AB, Wang J, Seffren V, Wiegand RE, Griffing S, Zell E. Public health impact of covid-19 vaccines in the US: observational study. BMJ 2022; : e069317.

12 Perez-Arce F, Angrisani M, Bennett D, Darling J, Kapteyn A, Thomas K. COVID-19 vaccines and mental distress. PLoS ONE 2021; 16: e0256406.

13 Koltai J, Raifman J, Bor J, McKee M, Stuckler D. COVID-19 Vaccination and Mental Health: A Difference-In-Difference Analysis of the Understanding America Study. American Journal of Preventive Medicine 2022; 62: 679–87.

14 Watkinson RE, Williams R, Gillibrand S, Sanders C, Sutton M. Ethnic inequalities in COVID-19 vaccine uptake and comparison to seasonal influenza vaccine uptake in Greater Manchester, UK: A cohort study. PLoS Med 2022; 19: e1003932.

15 Nafilyan V, Dolby T, Razieh C, et al. Sociodemographic inequality in COVID-19 vaccination coverage among elderly adults in England: a national linked data study. BMJ Open 2021; 11: e053402.

16 Gomez JMD, Du-Fay-de-Lavallaz JM, Fugar S, et al. Sex Differences in COVID-19 Hospitalization and Mortality. Journal of Women’s Health 2021; 30: 646–53.

17 Agyemang C, Richters A, Jolani S, et al. Ethnic minority status as social determinant for COVID-19 infection, hospitalisation, severity, ICU admission and deaths in the early phase of the pandemic: a meta-analysis. BMJ Glob Health 2021; 6: e007433.

18 Lo C-H, Nguyen LH, Drew DA, et al. Race, ethnicity, community-level socioeconomic factors, and risk of COVID-19 in the United States and the United Kingdom. eClinicalMedicine 2021; 38: 101029.

19 UKHSA. Omicron daily overview: 17 December 2021. https://assets.publishing.service.gov.uk/government/uploads/system/uploads/attachment_data/file/10421 00/20211217_OS_Daily_Omicron_Overview.pdf.

20 Curtis HJ, Inglesby P, Morton CE, et al. Trends and clinical characteristics of COVID-19 vaccine recipients: a federated analysis of 57.9 million patients’ primary care records in situ using OpenSAFELY. Br J Gen Pract 2022; 72: e51–62.

21 Tenforde MW, Self WH, Adams K, et al. Association Between mRNA Vaccination and COVID-19 Hospitalization and Disease Severity. JAMA 2021; 326: 2043.

22 Nguyen M. The Psychological Benefits of COVID-19 Vaccination. Advances in Public Health 2021; 2021: e1718800.

23 Nagarajan R, Krishnamoorthy Y, Basavarachar V, Dakshinamoorthy R. Prevalence of post-traumatic stress disorder among survivors of severe COVID-19 infections: A systematic review and meta-analysis. Journal of Affective Disorders 2022; 299: 52–9.

24 Bourmistrova NW, Solomon T, Braude P, Strawbridge R, Carter B. Long-term effects of COVID-19 on mental health: A systematic review. J Affect Disord 2022; 299: 118–25.

25 Zawilska JB, Kuczyńska K. Psychiatric and neurological complications of long COVID. Journal of Psychiatric Research 2022; 156: 349–60.

26 Kirchberger I, Meisinger C, Warm TD, Hyhlik-Dürr A, Linseisen J, Goßlau Y. Longitudinal course and predictors of health-related quality of life, mental health, and fatigue, in non-hospitalized individuals with or without post COVID-19 syndrome. In Review, 2023 DOI:10.21203/rs.3.rs-3221088/v1.

27 Pijls BG, Jolani S, Atherley A, et al. Demographic risk factors for COVID-19 infection, severity, ICU admission and death: a meta-analysis of 59 studies. BMJ Open 2021; 11: e044640.

28 Patel K, Robertson E, Kwong ASF, et al. Psychological Distress Before and During the COVID-19 Pandemic Among Adults in the United Kingdom Based on Coordinated Analyses of 11 Longitudinal Studies. JAMA Netw Open 2022; 5: e227629.

29 Hassan L, Sawyer C, Peek N, et al. COVID-19 vaccination uptake in people with severe mental illness: a UK-based cohort study. World Psychiatry 2022; 21: 153–4.

30 Hassan L, Peek N, Lovell K, et al. Disparities in COVID-19 infection, hospitalisation and death in people with schizophrenia, bipolar disorder, and major depressive disorder: a cohort study of the UK Biobank. Mol Psychiatry 2022; 27: 1248–55.

