## Supplement for "Impact of COVID-19 on mental illness in vaccinated and unvaccinated people: a population-based cohort study in OpenSAFELY"

**MANUSCRIPT TITLE**

Supplementary Table 6: Maximally adjusted hazard ratios and 95% CIs for mental illness events following diagnosis of COVID-19 in the pre-vaccination, vaccinated and unvaccinated cohorts, in people with no prior history of the event. 12

### SUPPLEMENTARY METHODS

#### Absolute excess risk

The absolute excess risk analysis was performed for each outcome in each cohort using the hazard ratio from the main analysis. To compare the outcomes across the cohorts, each of which have different lengths of follow-up, we calculated the absolute excess risk at 28 weeks. We accounted for age and sex in the analysis by using event counts and person days from eight subgroups defined by age and sex. They were females, aged 18-39; females, aged 40-59; females, aged 60-79; females, aged 80-110; males, aged 18-39; males, aged 40-59; males, aged 60-79; and males, aged 80-110). For each group, we calculated the average daily incidence of the outcome in the unexposed and the cumulative risk over time. We then used the relevant hazard ratio for that day (e.g., days 0 to 27 will have the coefficient for the term 'days0_28', while days 28 to 196 will have the coefficient for the term 'days28_197') to predict the expected cumulative survival in the exposed. Finally, we calculated the daily excess risk as the difference in cumulative survival for the unexposed and the expected cumulative survival in the exposed. The overall absolute excess risk was estimated using a weighted sum of the excess risks in each group, weighted by the proportions of individuals in age and sex strata in the pre-vaccination cohort.

#### Information governance

NHS England is the data controller of the NHS England OpenSAFELY COVID-19 Service; TPP is the data processor; all study authors using OpenSAFELY have the approval of NHS England. This implementation of OpenSAFELY is hosted within the TPP environment which is accredited to the ISO 27001 information security standard and is NHS IG Toolkit compliant.

Patient data has been pseudonymised for analysis and linkage using industry standard cryptographic hashing techniques; all pseudonymised datasets transmitted for linkage onto OpenSAFELY are encrypted; access to the NHS England OpenSAFELY COVID-19 service is via a virtual private network (VPN) connection; the researchers hold contracts with NHS England and only access the platform to initiate database queries and statistical models; all database activity is logged; only aggregate statistical outputs leave the platform environment following best practice for anonymisation of results such as statistical disclosure control for low cell counts.

The service adheres to the obligations of the UK General Data Protection Regulation (UK GDPR) and the Data Protection Act 2018. The service previously operated under notices initially issued in February 2020 by the the Secretary of State under Regulation 3(4) of the Health Service (Control of Patient Information) Regulations 2002 (COPI Regulations), which required organisations to process confidential patient information for COVID-19 purposes; this set aside the requirement for patient consent. As of 1 July 2023, the Secretary of State has requested that NHS England continue to operate the Service under the COVID-19 Directions 2020. In some cases of data sharing, the common law duty of confidence is met using, for example, patient consent or support from the Health Research Authority Confidentiality Advisory Group.

Taken together, these provide the legal bases to link patient datasets using the service. GP practices, which provide access to the primary care data, are required to share relevant health information to support the public health response to the pandemic, and have been informed of how the service operates.

This study was approved by the Health Research Authority [REC reference 22/PR/0095] and by the University of Bristol's Faculty of Health Sciences Ethics Committee [reference 117269].

### SUPPLEMENTARY TABLES

**Supplementary Table 1: Summary of covariate definitions.**

| Covariate | Type | Definition |
|---|---|---|
| Age | Continuous | Modelled as age in years using a restricted cubic spline with three knots at the 10th, 50th and 90th percentiles |
| Sex | Categorical | Male, Female |
| Ethnicity | Categorical | 1: White<br>2: Mixed<br>3: South Asian<br>4: Black<br>5: Other<br>Missing |
| Deprivation | Categorical | 10 categories from Index of Multiple Deprivation 2019 |
| Region | Categorical | East of England<br>London<br>Midlands<br>North East and Yorkshire<br>North West<br>South East<br>South West<br>Scotland<br>Wales |
| Smoking status | Categorical | E: Ever smoker<br>M: Missing<br>N: Never smoker<br>S: Current smoker |
| Care home status | Binary | 1 if care home resident; 0 otherwise |
| Consultation rate | Continuous | Number of GP consultations in 2019 |
| Health care worker | Binary | 1 if healthcare worker; 0 otherwise |
| Dementia | Binary | 1 if diagnosis present; 0 otherwise |
| Liver disease | Binary | 1 if diagnosis present; 0 otherwise |
| Chronic kidney disease | Binary | 1 if diagnosis present; 0 otherwise |
| Cancer | Categorical | 1 if diagnosis present; 0 otherwise |
| Hypertension | Binary | 1 if diagnosis present; 0 otherwise |
| Diabetes | Binary | 1 if diagnosis present; 0 otherwise |
| Obesity | Binary | 1 if BMI>=30 or coded diagnosis for obesity; 0 otherwise |

| | | |
|---|---|---|
| Chronic obstructive pulmonary disease (COPD) | Binary | 1 if diagnosis present; 0 otherwise |
| Acute myocardial infarction | Binary | 1 if diagnosis present; 0 otherwise |
| Ischaemic stroke | Binary | 1 if diagnosis present; 0 otherwise |
| Recent episode of depression | Binary | 1 if present ≤6 months prior to index date; 0 otherwise |
| History of depression | Binary | 1 if present >6 months prior to index data; 0 otherwise |
| Recent episode of anxiety | Binary | 1 if present ≤6 months prior to index date; 0 otherwise |
| History of anxiety | Binary | 1 if present >6 months prior to index date; 0 otherwise |
| Recent diagnosis of an eating disorder | Binary | 1 if present ≤6 months prior to index date; 0 otherwise |
| History of eating disorders | Binary | 1 if present >6 months prior to index date; 0 otherwise |
| Recent report of a serious mental illness | Binary | 1 if present ≤6 months prior to index date; 0 otherwise |
| History of serious mental illness | Binary | 1 if present >6 months prior to index date; 0 otherwise |
| Recent report of self-harm | Binary | 1 if present ≤6 months prior to index date; 0 otherwise |
| History of self-harm | Binary | 1 if present >6 months prior to index date; 0 otherwise |

**Supplementary Table 2: Summary of cohort characteristics.**

| Characteristic | Pre-vaccination cohort | Vaccinated cohort | Unvaccinated cohort |
|---|---|---|---|
| Start date | 01/01/2020, which is the approximate start date of the pandemic in the UK. | 01/06/2021, which is the date that the delta variant was thought to be ubiquitous in England. | 01/06/2021, which is the date that the delta variant was thought to be ubiquitous in England. |
| End date - exposure | 18/06/2021, which is the date when the Joint Committee for Vaccination and Immunisation (JCVI) phase 2, group 12 (all adults aged 18 years and older) become eligible for a COVID-19 vaccination. | 14/12/2021, which is the day that the UK Health Security Agency stated that over half of English cases they sampled have S Gene Target Failure, meaning they were likely Omicron, in this report. | 14/12/2021, which is the day that the UK Health Security Agency stated that over half of English cases they sampled have S Gene Target Failure, meaning they were likely Omicron, in this report. |
| End date - outcome | 14/12/2021, which is the day that the UK Health Security Agency stated that over half of English cases they sampled have S Gene Target Failure, meaning they were likely Omicron, in this report. | 14/12/2021, which is the day that the UK Health Security Agency stated that over half of English cases they sampled have S Gene Target Failure, meaning they were likely Omicron, in this report. | 14/12/2021, which is the day that the UK Health Security Agency stated that over half of English cases they sampled have S Gene Target Failure, meaning they were likely Omicron, in this report. |
| Exclusion criteria | Patients will be excluded if they meet any of the following criteria:<br>COVID-19 diagnosis recorded prior to their index date | Patients will be excluded if they meet any of the following criteria:<br>COVID-19 diagnosis recorded prior to their index date [Note: these individuals are required for a sensitivity analysis and so should not be removed at the data extraction stage]<br>They do not have a record of two vaccination doses prior to the study end date<br>They received a vaccination prior to 08-12-2020 (i.e., the start of the vaccination program)<br>They received a second dose vaccination before their first dose vaccination<br>They received a second dose vaccination less than three weeks after their first dose<br>They received mixed vaccine products before 07-05-2021 | Patients will be excluded if they meet any of the following criteria:<br>COVID-19 diagnosis recorded prior to their index date [Note: these individuals are required for a sensitivity analysis and so should not be removed at the data extraction stage]<br>They have a record of one or more vaccination doses prior to their index date<br>They could not be assigned to a vaccination group as defined by the Joint Committee on Vaccination and Immunisation (JCVI) |
| Follow-up start | Study start date. | Follow-up will start at the latest of the following dates (i.e., an individual's index date): Two | Follow-up will start at the latest of the following dates (i.e., an individual's index date): |

| | | | |
|---|---|---|---|
| | | weeks after their second vaccination; Study start date. | 12 weeks after they became eligible for vaccination; Study start date. |
| Follow-up end for exposure | Follow-up will end at the earliest of the following dates: Death; Outcome event; Study end date exposure; Deregistration date; Vaccination; Date when eligible for vaccination according to JCVI priority groupings. | Follow-up will end at the earliest of the following dates: Death; Outcome event; Study end date exposure; Deregistration date. | Follow-up will end at the earliest of the following dates: Death; Outcome event; Study end date exposure; Deregistration date; Vaccination. |
| Follow-up end for outcomes | Follow-up will end at the earliest of the following dates: Death; Outcome event; Study end date outcome; Deregistration date. | Follow-up will end at the earliest of the following dates: Death; Outcome event; Study end date outcome; Deregistration date. | Follow-up will end at the earliest of the following dates: Death; Outcome event; Study end date outcome; Deregistration date. |
| Cox regression time periods | [0,28), [28,197), [197, 365), [365,714) | [0,28), [28,197) | [0,28), [28,197) |

**Supplementary Table 3: Medical history of people in the pre-vaccination, vaccinated and unvaccinated cohorts.**

| Characteristic | | Pre-vaccination cohort | | Vaccinated cohort | | Unvaccinated cohort | |
|---|---|---|---|---|---|---|---|
| | | N (%) | COVID-19 diagnoses | N (%) | COVID-19 diagnoses | N (%) | COVID-19 diagnoses |
| All | | 17619987 | 975429 | 13716225 | 850023 | 3130581 | 147315 |
| GP visits in 2019 | 0 | 4738383 (26.9%) | 209559 | 3301473 (24.1%) | 190449 | 1685847 (53.9%) | 45657 |
| | 1-5 | 6527133 (37%) | 383793 | 5262447 (38.4%) | 344607 | 865647 (27.7%) | 55965 |
| | 6+ | 6354465 (36.1%) | 382071 | 5152299 (37.6%) | 314967 | 579087 (18.5%) | 45699 |
| History of depression | None | 12540909 (71.2%) | 678621 | 9506259 (69.3%) | 569673 | 2424975 (77.5%) | 99657 |
| | More than six months ago | 4629915 (26.3%) | 265431 | 3908361 (28.5%) | 259293 | 647697 (20.7%) | 43281 |
| | Within six months | 449157 (2.5%) | 31371 | 301599 (2.2%) | 21057 | 57909 (1.8%) | 4377 |
| History of eating disorders | None | 17442471 (99%) | 964419 | 13570203 (98.9%) | 839469 | 3100647 (99%) | 145191 |
| | More than six months ago | 171861 (1%) | 10611 | 140967 (1%) | 10221 | 28803 (0.9%) | 2037 |
| | Within six months | 5649 (0%) | 393 | 5055 (0%) | 333 | 1131 (0%) | 87 |
| History of general anxiety | None | 13940733 (79.1%) | 757449 | 10593231 (77.2%) | 639285 | 2578311 (82.4%) | 109329 |
| | More than six months ago | 3378171 (19.2%) | 196617 | 2913429 (21.2%) | 195711 | 507069 (16.2%) | 34401 |
| | Within six months | 301083 (1.7%) | 21363 | 209559 (1.5%) | 15027 | 45201 (1.4%) | 3585 |
| History of self-harm | None | 16849539 (95.6%) | 930099 | 13130817 (95.7%) | 813273 | 2969463 (94.9%) | 137529 |
| | More than six months ago | 744843 (4.2%) | 43503 | 569349 (4.2%) | 35781 | 154821 (4.9%) | 9429 |
| | Within six months | 25605 (0.1%) | 1821 | 16053 (0.1%) | 963 | 6291 (0.2%) | 357 |
| History of serious mental illness | None | 15097599 (85.7%) | 828747 | 11647629 (84.9%) | 714519 | 2753379 (88%) | 123141 |
| | More than six months ago | 2391747 (13.6%) | 137529 | 1990077 (14.5%) | 130365 | 358479 (11.5%) | 22941 |
| | Within six months | 130641 (0.7%) | 9153 | 78513 (0.6%) | 5133 | 18723 (0.6%) | 1239 |
| Medical history | Acute myocardial infarction | 449439 (2.6%) | 22239 | 417135 (3%) | 16527 | 21087 (0.7%) | 1131 |
| | Cancer | 5273097 (29.9%) | 327495 | 4684545 (34.2%) | 357519 | 599169 (19.1%) | 49731 |
| | Chronic kidney disease | 1126839 (6.4%) | 54903 | 1044879 (7.6%) | 37965 | 46425 (1.5%) | 2649 |
| | COPD | 586821 (3.3%) | 27807 | 530235 (3.9%) | 20667 | 28299 (0.9%) | 1395 |
| | Dementia | 211965 (1.2%) | 21621 | 170955 (1.2%) | 6285 | 5895 (0.2%) | 333 |
| | Diabetes | 1588881 (9%) | 94269 | 1458759 (10.6%) | 78333 | 118965 (3.8%) | 7911 |
| | Hypertension | 6135309 (34.8%) | 309891 | 5462565 (39.8%) | 283599 | 477465 (15.3%) | 31245 |
| | Liver disease | 128733 (0.7%) | 6849 | 114603 (0.8%) | 5673 | 15849 (0.5%) | 735 |
| | Obesity | 4385937 (24.9%) | 279231 | 3786285 (27.6%) | 259983 | 468033 (15%) | 33099 |
| | Stroke | 285837 (1.6%) | 15123 | 265863 (1.9%) | 9633 | 13425 (0.4%) | 729 |

COPD: chronic obstructive pulmonary disease.

**Supplementary Table 4: Maximally adjusted hazard ratios and 95% CIs for mental illness events following hospitalised COVID-19 in the pre-vaccination, vaccinated and unvaccinated cohorts.**

| Outcome | Time since COVID-19 | Pre-vaccination cohort | Vaccinated cohort | Unvaccinated cohort |
|---|---|---|---|---|
| Depression | Day 0 | 265 (253-278) | 129 (118-142) | 273 (248-300) |
| | 1-4 weeks | 16.5 (15.8-17.3) | 12.9 (12.0-14.0) | 15.6 (14.1-17.2) |
| | 5-28 weeks | 1.82 (1.72-1.93) | 1.54 (1.31-1.81) | 1.92 (1.58-2.34) |
| | 29-52 weeks | 1.45 (1.35-1.56) | - | - |
| | 53-102 weeks | 1.34 (1.21-1.48) | - | - |
| Serious mental illness | Day 0 | 227 (207-250) | 104 (83.4-129) | 144 (113-183) |
| | 1-4 weeks | 9.65 (8.71-10.7) | 6.38 (5.21-7.80) | 8.74 (6.97-11.0) |
| | 5-28 weeks | 1.55 (1.39-1.74) | 0.91 (0.62-1.34) | 1.01 (0.64-1.58) |
| | 29-52 weeks | 1.38 (1.22-1.57) | - | - |
| | 53-102 weeks | 1.03 (0.81-1.29) | - | - |
| General anxiety | Day 0 | 307 (290-324) | 152 (135-170) | 272 (242-306) |
| | 1-4 weeks | 21.3 (20.3-22.3) | 17.3 (15.9-18.8) | 27.2 (24.6-30.1) |
| | 5-28 weeks | 1.96 (1.83-2.10) | 1.62 (1.33-1.97) | 3.04 (2.56-3.61) |
| | 29-52 weeks | 1.42 (1.31-1.55) | - | - |
| | 53-102 weeks | 1.21 (1.07-1.37) | - | - |
| Post-traumatic stress disorder | Day 0 | 403 (309-526) | 285 (179-452) | 372 (250-556) |
| | 1-4 weeks | 19.9 (15.6-25.5) | 26.3 (19.3-35.8) | 14.0 (8.40-23.4) |
| | 5-28 weeks | 4.11 (3.30-5.13) | 2.10 (0.96-4.59) | 2.67 (1.36-5.22) |
| | 29-52 weeks | 2.30 (1.71-3.10) | - | - |
| | 53-102 weeks | 1.97 (1.17-3.32) | - | - |
| Eating disorders | Day 0 | 172 (97.1-304) | † | † |
| | 1-4 weeks | 9.08 (5.37-15.3) | † | † |
| | 5-28 weeks | 1.68 (1.01-2.79) | † | † |
| | 29-52 weeks | 1.34 (0.73-2.46) | - | - |
| | 53-102 weeks | 1.94 (0.96-3.90) | - | - |
| Addiction | Day 0 | 285 (203-400) | † | 366 (260-515) |
| | 1-4 weeks | 8.45 (5.54-12.9) | † | 8.71 (5.36-14.1) |
| | 5-28 weeks | 1.48 (0.95-2.32) | † | 0.82 (0.26-2.53) |
| | 29-52 weeks | 1.65 (1.01-2.69) | - | - |
| | 53-102 weeks | 1.25 (0.52-3.02) | - | - |
| Self-harm | Day 0 | 80.5 (55.5-117) | † | † |
| | 1-4 weeks | 3.58 (2.49-5.13) | † | † |
| | 5-28 weeks | 1.78 (1.43-2.22) | † | † |
| | 29-52 weeks | 1.67 (1.29-2.15) | - | - |
| | 53-102 weeks | 1.87 (1.34-2.61) | - | - |

† Insufficient events for estimation

**Supplementary Table 5: Maximally adjusted hazard ratios and 95% Cis for mental illness events following non-hospitalised COVID-19 in the pre-vaccination, vaccinated and unvaccinated cohorts.**

| Outcome | Time since COVID-19 | Pre-vaccination cohort | Vaccinated cohort | Unvaccinated cohort |
|---|---|---|---|---|
| Depression | Day 0 | 17.7 (16.9-18.5) | 5.41 (5.03-5.82) | 7.79 (6.79-8.95) |
| | 1-4 weeks | 1.22 (1.17-1.26) | 0.92 (0.88-0.95) | 1.11 (1.02-1.20) |
| | 5-28 weeks | 1.22 (1.20-1.24) | 1.10 (1.07-1.13) | 1.26 (1.19-1.33) |
| | 29-52 weeks | 1.15 (1.13-1.17) | - | - |
| | 53-102 weeks | 1.15 (1.12-1.19) | - | - |
| Serious mental illness | Day 0 | 16.9 (15.5-18.4) | 5.36 (4.63-6.21) | 11.1 (8.90-13.9) |
| | 1-4 weeks | 1.04 (0.97-1.11) | 0.79 (0.73-0.86) | 0.98 (0.83-1.15) |
| | 5-28 weeks | 1.18 (1.15-1.21) | 1.07 (1.01-1.12) | 1.15 (1.03-1.28) |
| | 29-52 weeks | 1.14 (1.10-1.17) | - | - |
| | 53-102 weeks | 1.14 (1.08-1.21) | - | - |
| General anxiety | Day 0 | 18.8 (17.9-19.8) | 5.47 (5.01-5.97) | 9.44 (8.19-10.9) |
| | 1-4 weeks | 1.38 (1.33-1.44) | 0.97 (0.93-1.01) | 1.25 (1.15-1.36) |
| | 5-28 weeks | 1.23 (1.21-1.25) | 1.11 (1.08-1.15) | 1.31 (1.22-1.39) |
| | 29-52 weeks | 1.12 (1.10-1.15) | - | - |
| | 53-102 weeks | 1.14 (1.10-1.18) | - | - |
| Post-traumatic stress disorder | Day 0 | 21.6 (16.9-27.6) | 7.00 (4.72-10.4) | 13.8 (8.75-21.7) |
| | 1-4 weeks | 0.99 (0.79-1.24) | 0.61 (0.46-0.81) | 0.70 (0.46-1.09) |
| | 5-28 weeks | 0.99 (0.91-1.08) | 1.01 (0.86-1.19) | 0.97 (0.73-1.29) |
| | 29-52 weeks | 1.00 (0.91-1.10) | - | - |
| | 53-102 weeks | 1.07 (0.90-1.28) | - | - |
| Eating disorders | Day 0 | 19.1 (13.7-26.8) | 4.70 (2.64-8.37) | † |
| | 1-4 weeks | 1.52 (1.20-1.92) | 0.74 (0.53-1.05) | † |
| | 5-28 weeks | 1.24 (1.11-1.39) | 0.87 (0.68-1.12) | † |
| | 29-52 weeks | 1.08 (0.95-1.23) | - | - |
| | 53-102 weeks | 1.14 (0.89-1.47) | - | - |
| Addiction | Day 0 | 48.1 (39.9-58.0) | 12.5 (8.35-18.7) | 22.9 (16.1-32.7) |
| | 1-4 weeks | 0.97 (0.75-1.25) | 0.70 (0.49-1.01) | 0.53 (0.32-0.87) |
| | 5-28 weeks | 0.90 (0.81-1.00) | 0.63 (0.47-0.83) | 0.77 (0.56-1.06) |
| | 29-52 weeks | 0.94 (0.84-1.06) | - | - |
| | 53-102 weeks | 0.86 (0.67-1.12) | - | - |
| Self-harm | Day 0 | 19.3 (16.3-22.8) | 10.1 (7.93-13.0) | 10.4 (6.82-16.0) |
| | 1-4 weeks | 1.10 (0.96-1.27) | 0.94 (0.79-1.11) | 1.08 (0.81-1.44) |
| | 5-28 weeks | 1.13 (1.07-1.20) | 1.11 (0.99-1.25) | 0.93 (0.74-1.18) |
| | 29-52 weeks | 1.15 (1.08-1.23) | - | - |
| | 53-102 weeks | 1.09 (0.95-1.24) | - | - |
| Suicide | Day 0 | 12.0 (4.34-33.4) | † | † |
| | 1-4 weeks | 2.38 (1.40-4.04) | † | † |
| | 5-28 weeks | 0.82 (0.57-1.17) | † | † |
| | 29-52 weeks | 0.80 (0.54-1.20) | - | - |
| | 53-102 weeks | 0.69 (0.29-1.66) | - | - |

† Insufficient events for estimation

**Supplementary Table 6: Maximally adjusted hazard ratios and 95% CIs for mental illness events following diagnosis of COVID-19 in the pre-vaccination, vaccinated and unvaccinated cohorts, in people with no prior history of the event.**

| Outcome | Time since COVID-19 | Pre-vaccination cohort | Vaccinated cohort | Unvaccinated cohort |
|---|---|---|---|---|
| Depression | Day 0 | 18.1 (16.7-19.6) | 5.94 (5.12-6.89) | 11.3 (8.81-14.4) |
| | 1-4 weeks | 1.58 (1.50-1.) | 1.11 (1.03-1.19) | 1.44 (1.23-1.67) |
| | 5-28 weeks | 1.40 (1.37-1.44) | 1.26 (1.20-1.32) | 1.48 (1.32-1.67) |
| | 29-52 weeks | 1.23 (1.20-1.27) | - | - |
| | 53-102 weeks | 1.30 (1.24-1.37) | - | - |
| Serious mental illness | Day 0 | 11.0 (9.62-12.7) | 4.20 (3.25-5.43) | 8.84 (6.15-12.7) |
| | 1-4 weeks | 1.18 (1.08-1.29) | 0.94 (0.83-1.06) | 1.20 (0.96-1.50) |
| | 5-28 weeks | 1.31 (1.26-1.35) | 1.23 (1.14-1.32) | 1.24 (1.05-1.47) |
| | 29-52 weeks | 1.24 (1.19-1.29) | - | - |
| | 53-102 weeks | 1.21 (1.12-1.31) | - | - |

† Insufficient events for estimation

**Supplementary Table 7: Maximally adjusted hazard ratios and 95% CIs for mental illness events following diagnosis of COVID-19 in the pre-vaccination, vaccinated and unvaccinated cohorts, in people with prior history of event, more than six months ago.**

| Outcome | Time since COVID-19 | Pre-vaccination cohort | Vaccinated cohort | Unvaccinated cohort |
|---|---|---|---|---|
| Depression | Day 0 | 41.0 (39.5-42.7) | 10.7 (10.0-11.4) | 24.4 (22.1-27.1) |
| | 1-4 weeks | 2.16 (2.08-2.23) | 1.22 (1.17-1.27) | 2.06 (1.91-2.23) |
| | 5-28 weeks | 1.17 (1.15-1.19) | 1.11 (1.07-1.14) | 1.29 (1.20-1.39) |
| | 29-52 weeks | 1.12 (1.10-1.15) | - | - |
| | 53-102 weeks | 1.08 (1.04-1.13) | - | - |
| Serious mental illness | Day 0 | 47.7 (44.2-51.4) | 11.2 (9.70-12.9) | 27.4 (22.4-33.5) |
| | 1-4 weeks | 1.79 (1.65-1.94) | 0.89 (0.80-1.00) | 1.56 (1.30-1.88) |
| | 5-28 weeks | 1.09 (1.04-1.13) | 1.00 (0.93-1.08) | 1.01 (0.86-1.19) |
| | 29-52 weeks | 1.07 (1.02-1.12) | - | - |
| | 53-102 weeks | 1.09 (0.99-1.19) | - | - |

† Insufficient events for estimation

**Supplementary Table 8: Maximally adjusted hazard ratios and 95% CIs for mental illness events following diagnosis of COVID-19 in the pre-vaccination, vaccinated and unvaccinated cohorts, in people with prior history of event, within six months.**

| Outcome | Time since COVID-19 | Pre-vaccination cohort | Vaccinated cohort | Unvaccinated cohort |
|---|---|---|---|---|
| Depression | Day 0 | 39.3 (36.4-42.4) | 9.66 (8.62-10.8) | 12.3 (9.97-15.1) |
| | 1-4 weeks | 1.66 (1.52-1.80) | 1.01 (0.93-1.10) | 1.33 (1.15-1.54) |
| | 5-28 weeks | 1.07 (1.02-1.12) | 0.94 (0.87-1.00) | 1.01 (0.88-1.16) |
| | 29-52 weeks | 1.02 (0.97-1.08) | - | - |
| | 53-102 weeks | 1.11 (1.01-1.23) | - | - |
| Serious mental illness | Day 0 | 65.1 (56.8-74.7) | 14.4 (11.4-18.3) | 19.9 (13.1-30.0) |
| | 1-4 weeks | 1.88 (1.57-2.26) | 1.07 (0.87-1.33) | 1.34 (0.94-1.93) |
| | 5-28 weeks | 1.02 (0.92-1.14) | 0.93 (0.78-1.10) | 1.13 (0.83-1.52) |
| | 29-52 weeks | 0.87 (0.76-1.00) | - | - |
| | 53-102 weeks | 1.02 (0.80-1.30) | - | - |

† Insufficient events for estimation

**Supplementary Table 9: Maximally adjusted hazard ratios and 95% CIs for mental illness events following diagnosis of COVID-19 in the pre-vaccination, vaccinated and unvaccinated cohorts, in people with history of COVID-19.**

| Outcome | Time since COVID-19 | Vaccinated cohort | Unvaccinated cohort |
|---|---|---|---|
| Depression | Day 0 | 41.4 (34.0-50.4) | 51.4 (35.2-75.1) |
| | 1-4 weeks | 1.30 (0.99-1.72) | 2.58 (1.73-3.85) |
| | 5-28 weeks | 1.13 (0.94-1.35) | 1.68 (1.20-2.35) |
| Serious mental illness | Day 0 | 45.1 (30.2-67.4) | † |
| | 1-4 weeks | 0.82 (0.41-1.64) | † |
| | 5-28 weeks | 0.87 (0.58-1.31) | † |

† Insufficient events for estimation

**Supplementary Table 10: Maximally adjusted hazard ratios and 95% CIs for mental illness events following diagnosis of COVID-19 in the pre-vaccination, vaccinated and unvaccinated cohorts for age group 18-39.**

| Outcome | Time since COVID-19 | Pre-vaccination cohort | Vaccinated cohort | Unvaccinated cohort |
|---|---|---|---|---|
| Depression | Day 0 | 6.10 (5.47-6.82) | 2.15 (1.77-2.61) | 7.19 (6.08-8.50) |
| | 1-4 weeks | 1.07 (1.01-1.12) | 0.88 (0.82-0.94) | 1.30 (1.19-1.42) |
| | 5-28 weeks | 1.19 (1.16-1.21) | 1.06 (1.02-1.11) | 1.23 (1.14-1.32) |
| | 29-52 weeks | 1.16 (1.14-1.19) | - | - |
| | 53-102 weeks | 1.19 (1.13-1.25) | - | - |
| Serious mental illness | Day 0 | 5.60 (4.58-6.84) | 1.85 (1.24-2.76) | 7.91 (5.80-10.8) |
| | 1-4 weeks | 0.97 (0.88-1.07) | 0.83 (0.72-0.94) | 1.05 (0.87-1.26) |
| | 5-28 weeks | 1.13 (1.09-1.17) | 1.10 (1.01-1.19) | 1.15 (0.99-1.33) |
| | 29-52 weeks | 1.13 (1.08-1.17) | - | - |
| | 53-102 weeks | 1.05 (0.96-1.15) | - | - |

† Insufficient events for estimation

**Supplementary Table 11: Maximally adjusted hazard ratios and 95% CIs for mental illness events following diagnosis of COVID-19 in the pre-vaccination, vaccinated and unvaccinated cohorts for age group 40-59.**

| Outcome | Time since COVID-19 | Pre-vaccination cohort | Vaccinated cohort | Unvaccinated cohort |
|---|---|---|---|---|
| Depression | Day 0 | 24.6 (23.0-26.2) | 4.52 (4.01-5.10) | 30.0 (26.1-34.4) |
| | 1-4 weeks | 2.11 (2.01-2.20) | 1.10 (1.05-1.16) | 2.40 (2.15-2.68) |
| | 5-28 weeks | 1.22 (1.19-1.25) | 1.13 (1.08-1.17) | 1.39 (1.26-1.54) |
| | 29-52 weeks | 1.16 (1.13-1.19) | - | - |
| | 53-102 weeks | 1.21 (1.15-1.27) | - | - |
| Serious mental illness | Day 0 | 22.4 (19.8-25.5) | 4.46 (3.51-5.67) | 24.3 (18.4-32.2) |
| | 1-4 weeks | 1.44 (1.30-1.59) | 0.80 (0.71-0.91) | 1.48 (1.16-1.89) |
| | 5-28 weeks | 1.12 (1.06-1.17) | 1.05 (0.97-1.14) | 1.11 (0.92-1.34) |
| | 29-52 weeks | 1.09 (1.04-1.15) | - | - |
| | 53-102 weeks | 1.23 (1.11-1.35) | - | - |

† Insufficient events for estimation

**Supplementary Table 12: Maximally adjusted hazard ratios and 95% CIs for mental illness events following diagnosis of COVID-19 in the pre-vaccination, vaccinated and unvaccinated cohorts for age group 60-79.**

| Outcome | Time since COVID-19 | Pre-vaccination cohort | Vaccinated cohort | Unvaccinated cohort |
|---|---|---|---|---|
| Depression | Day 0 | 111 (105-117) | 23.4 (21.3-25.7) | 477 (435-522) |
| | 1-4 weeks | 4.46 (4.21-4.71) | 1.67 (1.55-1.80) | 21.6 (19.5-23.9) |
| | 5-28 weeks | 1.44 (1.39-1.50) | 1.17 (1.09-1.24) | 2.79 (2.26-3.43) |
| | 29-52 weeks | 1.19 (1.14-1.24) | - | - |
| | 53-102 weeks | 1.11 (1.03-1.20) | - | - |
| Serious mental illness | Day 0 | 120 (109-132) | 27.2 (22.7-32.5) | 123 (86.7-175) |
| | 1-4 weeks | 3.31 (2.93-3.73) | 1.35 (1.13-1.61) | 6.29 (4.36-9.08) |
| | 5-28 weeks | 1.54 (1.43-1.66) | 1.06 (0.92-1.22) | 1.68 (1.03-2.75) |
| | 29-52 weeks | 1.31 (1.20-1.42) | - | - |
| | 53-102 weeks | 1.08 (0.93-1.26) | - | - |

† Insufficient events for estimation

**Supplementary Table 13: Maximally adjusted hazard ratios and 95% CIs for mental illness events following diagnosis of COVID-19 in the pre-vaccination, vaccinated and unvaccinated cohorts for age group 80-110.**

| Outcome | Time since COVID-19 | Pre-vaccination cohort | Vaccinated cohort | Unvaccinated cohort |
|---|---|---|---|---|
| Depression | Day 0 | 131 (122-142) | 71.3 (63.3-80.3) | 279 (196-397) |
| | 1-4 weeks | 3.22 (2.90-3.59) | 2.74 (2.40-3.14) | 6.75 (3.92-11.6) |
| | 5-28 weeks | 1.41 (1.31-1.51) | 1.27 (1.10-1.47) | 1.92 (0.90-4.08) |
| | 29-52 weeks | 1.11 (1.02-1.21) | - | - |
| | 53-102 weeks | 0.89 (0.80-1.01) | - | - |
| Serious mental illness | Day 0 | 103 (87.1-122) | 58.3 (43.7-77.6) | † |
| | 1-4 weeks | 3.60 (2.95-4.40) | 1.92 (1.36-2.71) | † |
| | 5-28 weeks | 1.73 (1.51-1.97) | 1.21 (0.89-1.65) | † |
| | 29-52 weeks | 1.42 (1.22-1.67) | - | - |
| | 53-102 weeks | 1.04 (0.84-1.29) | - | - |

† Insufficient events for estimation

**Supplementary Table 14: Maximally adjusted hazard ratios and 95% CIs for mental illness events following diagnosis of COVID-19 in the pre-vaccination, vaccinated and unvaccinated cohorts for women.**

| Outcome | Time since COVID-19 | Pre-vaccination cohort | Vaccinated cohort | Unvaccinated cohort |
|---|---|---|---|---|
| Depression | Day 0 | 26.4 (25.3-27.6) | 7.33 (6.80-7.91) | 17.2 (15.5-19.2) |
| | 1-4 weeks | 1.77 (1.71-1.84) | 1.11 (1.06-1.15) | 1.63 (1.51-1.76) |
| | 5-28 weeks | 1.23 (1.21-1.25) | 1.11 (1.08-1.15) | 1.25 (1.16-1.33) |
| | 29-52 weeks | 1.15 (1.13-1.17) | - | - |
| | 53-102 weeks | 1.16 (1.12-1.21) | - | - |
| Serious mental illness | Day 0 | 25.5 (23.4-27.8) | 6.18 (5.22-7.32) | 15.2 (12.0-19.2) |
| | 1-4 weeks | 1.34 (1.24-1.45) | 0.87 (0.79-0.96) | 1.16 (0.96-1.39) |
| | 5-28 weeks | 1.19 (1.15-1.23) | 1.11 (1.04-1.18) | 1.12 (0.97-1.29) |
| | 29-52 weeks | 1.14 (1.10-1.19) | - | - |
| | 53-102 weeks | 1.13 (1.05-1.21) | - | - |

† Insufficient events for estimation

**Supplementary Table 15: Maximally adjusted hazard ratios and 95% CIs for mental illness events following diagnosis of COVID-19 in the pre-vaccination, vaccinated and unvaccinated cohorts for men.**

| Outcome | Time since COVID-19 | Pre-vaccination cohort | Vaccinated cohort | Unvaccinated cohort |
|---|---|---|---|---|
| Depression | Day 0 | 53.5 (51.2-55.8) | 11.2 (10.2-12.2) | 24.0 (20.8-27.8) |
| | 1-4 weeks | 2.26 (2.16-2.36) | 1.29 (1.21-1.36) | 2.22 (1.99-2.47) |
| | 5-28 weeks | 1.27 (1.24-1.30) | 1.11 (1.06-1.16) | 1.40 (1.26-1.55) |
| | 29-52 weeks | 1.18 (1.15-1.21) | - | - |
| | 53-102 weeks | 1.18 (1.11-1.24) | - | - |
| Serious mental illness | Day 0 | 36.1 (33.0-39.6) | 10.4 (8.76-12.4) | 24.5 (19.4-30.9) |
| | 1-4 weeks | 1.70 (1.56-1.85) | 0.99 (0.88-1.12) | 1.78 (1.47-2.15) |
| | 5-28 weeks | 1.22 (1.17-1.27) | 1.02 (0.93-1.11) | 1.14 (0.95-1.36) |
| | 29-52 weeks | 1.16 (1.10-1.21) | - | - |
| | 53-102 weeks | 1.13 (1.02-1.24) | - | - |

† Insufficient events for estimation

**Supplementary Table 16: Maximally adjusted hazard ratios and 95% CIs for mental illness events following diagnosis of COVID-19 in the pre-vaccination, vaccinated and unvaccinated cohorts for White ethnicity.**

| Outcome | Time since COVID-19 | Pre-vaccination cohort | Vaccinated cohort | Unvaccinated cohort |
|---|---|---|---|---|
| Depression | Day 0 | 34.2 (33.0-35.4) | 8.67 (8.17-9.20) | 18.3 (16.7-20.2) |
| | 1-4 weeks | 1.94 (1.88-1.99) | 1.16 (1.12-1.20) | 1.74 (1.62-1.86) |
| | 5-28 weeks | 1.24 (1.22-1.26) | 1.11 (1.08-1.14) | 1.25 (1.18-1.33) |
| | 29-52 weeks | 1.15 (1.13-1.17) | - | - |
| | 53-102 weeks | 1.17 (1.13-1.21) | - | - |
| Serious mental illness | Day 0 | 29.2 (27.3-31.3) | 7.65 (6.73-8.69) | 17.7 (14.6-21.3) |
| | 1-4 weeks | 1.42 (1.33-1.51) | 0.91 (0.84-0.99) | 1.33 (1.15-1.54) |
| | 5-28 weeks | 1.21 (1.17-1.24) | 1.07 (1.02-1.13) | 1.12 (0.99-1.26) |
| | 29-52 weeks | 1.15 (1.11-1.18) | - | - |
| | 53-102 weeks | 1.13 (1.06-1.21) | - | - |

† Insufficient events for estimation

**Supplementary Table 17: Maximally adjusted hazard ratios and 95% CIs for mental illness events following diagnosis of COVID-19 in the pre-vaccination, vaccinated and unvaccinated cohorts for South Asian ethnicity.**

| Outcome | Time since COVID-19 | Pre-vaccination cohort | Vaccinated cohort | Unvaccinated cohort |
|---|---|---|---|---|
| Depression | Day 0 | 29.2 (25.5-33.4) | 12.4 (9.36-16.5) | 36.6 (27.0-49.6) |
| | 1-4 weeks | 1.99 (1.80-2.21) | 1.12 (0.91-1.37) | 2.13 (1.63-2.80) |
| | 5-28 weeks | 1.33 (1.27-1.41) | 1.15 (0.99-1.32) | 1.40 (1.09-1.79) |
| | 29-52 weeks | 1.23 (1.16-1.30) | - | - |
| | 53-102 weeks | 1.14 (1.02-1.27) | - | - |
| Serious mental illness | Day 0 | 34.8 (28.3-42.6) | 9.31 (5.29-16.4) | † |
| | 1-4 weeks | 1.68 (1.40-2.03) | 0.88 (0.59-1.31) | † |
| | 5-28 weeks | 1.13 (1.03-1.24) | 0.97 (0.74-1.27) | † |
| | 29-52 weeks | 1.09 (0.98-1.21) | - | - |
| | 53-102 weeks | 0.99 (0.81-1.20) | - | - |

† Insufficient events for estimation

**Supplementary Table 18: Maximally adjusted hazard ratios and 95% CIs for mental illness events following diagnosis of COVID-19 in the pre-vaccination, vaccinated and unvaccinated cohorts for Black ethnicity.**

| Outcome | Time since COVID-19 | Pre-vaccination cohort | Vaccinated cohort | Unvaccinated cohort |
|---|---|---|---|---|
| Depression | Day 0 | 36.8 (28.5-47.4) | 9.63 (5.16-18.0) | 18.0 (11.1-29.0) |
| | 1-4 weeks | 1.93 (1.55-2.41) | 1.65 (1.16-2.33) | 1.88 (1.34-2.64) |
| | 5-28 weeks | 1.09 (0.97-1.23) | 1.21 (0.89-1.63) | 1.36 (1.03-1.78) |
| | 29-52 weeks | 1.22 (1.08-1.38) | - | - |
| | 53-102 weeks | 0.86 (0.65-1.13) | - | - |
| Serious mental illness | Day 0 | 49.8 (35.8-69.2) | † | † |
| | 1-4 weeks | 2.46 (1.82-3.32) | † | † |
| | 5-28 weeks | 0.94 (0.77-1.14) | † | † |
| | 29-52 weeks | 1.18 (0.97-1.44) | - | - |
| | 53-102 weeks | 1.13 (0.77-1.67) | - | - |

† Insufficient events for estimation

**Supplementary Table 19: Maximally adjusted hazard ratios and 95% CIs for mental illness events following diagnosis of COVID-19 in the pre-vaccination, vaccinated and unvaccinated cohorts for Other ethnicity.**

| Outcome | Time since COVID-19 | Pre-vaccination cohort | Vaccinated cohort | Unvaccinated cohort |
|---|---|---|---|---|
| Depression | Day 0 | 35.8 (26.7-48.1) | 3.42 (1.29-9.09) | 25.5 (13.4-48.5) |
| | 1-4 weeks | 2.52 (2.01-3.16) | 1.35 (0.95-1.93) | 2.41 (1.47-3.93) |
| | 5-28 weeks | 1.56 (1.39-1.76) | 1.18 (0.89-1.57) | 1.86 (1.20-2.90) |
| | 29-52 weeks | 1.39 (1.21-1.59) | - | - |
| | 53-102 weeks | 1.31 (0.99-1.72) | - | - |
| Serious mental illness | Day 0 | 26.3 (15.2-45.6) | † | † |
| | 1-4 weeks | 1.48 (0.93-2.37) | † | † |
| | 5-28 weeks | 1.54 (1.27-1.87) | † | † |
| | 29-52 weeks | 1.53 (1.23-1.89) | - | - |
| | 53-102 weeks | 1.05 (0.64-1.72) | - | - |

† Insufficient events for estimation

**Supplementary Table 20: Maximally adjusted hazard ratios and 95% CIs for mental illness events following diagnosis of COVID-19 in the pre-vaccination, vaccinated and unvaccinated cohorts for Mixed ethnicity.**

| Outcome | Time since COVID-19 | Pre-vaccination cohort | Vaccinated cohort | Unvaccinated cohort |
|---|---|---|---|---|
| Depression | Day 0 | 20.4 (14.1-29.4) | 4.73 (2.15-10.4) | 13.5 (7.39-24.6) |
| | 1-4 weeks | 1.46 (1.11-1.93) | 0.75 (0.49-1.17) | 2.08 (1.48-2.93) |
| | 5-28 weeks | 1.23 (1.08-1.39) | 0.81 (0.60-1.10) | 0.89 (0.61-1.32) |
| | 29-52 weeks | 1.10 (0.95-1.26) | - | - |
| | 53-102 weeks | 1.54 (1.20-1.96) | - | - |
| Serious mental illness | Day 0 | 23.5 (13.6-40.7) | † | † |
| | 1-4 weeks | 2.15 (1.50-3.09) | † | † |
| | 5-28 weeks | 1.03 (0.83-1.28) | † | † |
| | 29-52 weeks | 1.07 (0.85-1.34) | - | - |
| | 53-102 weeks | 1.66 (1.13-2.45) | - | - |

† Insufficient events for estimation

**Supplementary Table 21: Excess events per 100,000 people at 28 weeks post-COVID-19 in the pre-vaccination, vaccinated and unvaccinated cohorts.**

| Outcome | Pre-vaccination cohort | | | Vaccinated cohort | | | Unvaccinated cohort | | |
|---|---|---|---|---|---|---|---|---|---|
| | Day zero included | Day zero excluded | Difference (%) | Day zero included | Day zero excluded | Difference (%) | Day zero included | Day zero excluded | Difference (%) |
| Depression | 1020 | 691 | 32 | 449 | 337 | 25 | 1009 | 802 | 21 |
| Serious mental illness | 227 | 141 | 38 | 60 | 34 | 43 | 199 | 130 | 34 |
| General anxiety | 817 | 571 | 30 | 357 | 280 | 22 | 1012 | 808 | 20 |
| PTSD | 26 | 13 | 48 | 5 | 1 | 85 | 27 | 11 | 60 |
| Eating disorders | 15 | 11 | 29 | -4 | -5 | -32 | 11 | 7 | 34 |
| Addiction | 21 | 1 | 95 | -9 | -14 | -44 | 5 | -30 | 705 |
| Self-harm | 38 | 23 | 40 | 31 | 23 | 26 | -1 | -11 | -1985 |
| Suicide | 303 | 20 | 93 | † | † | † | † | † | † |

† Insufficient events for estimation; PTSD = post-traumatic stress disorder.

### SUPPLEMENTARY FIGURES

#### Supplementary Figure 1: COVID-19 cases over time

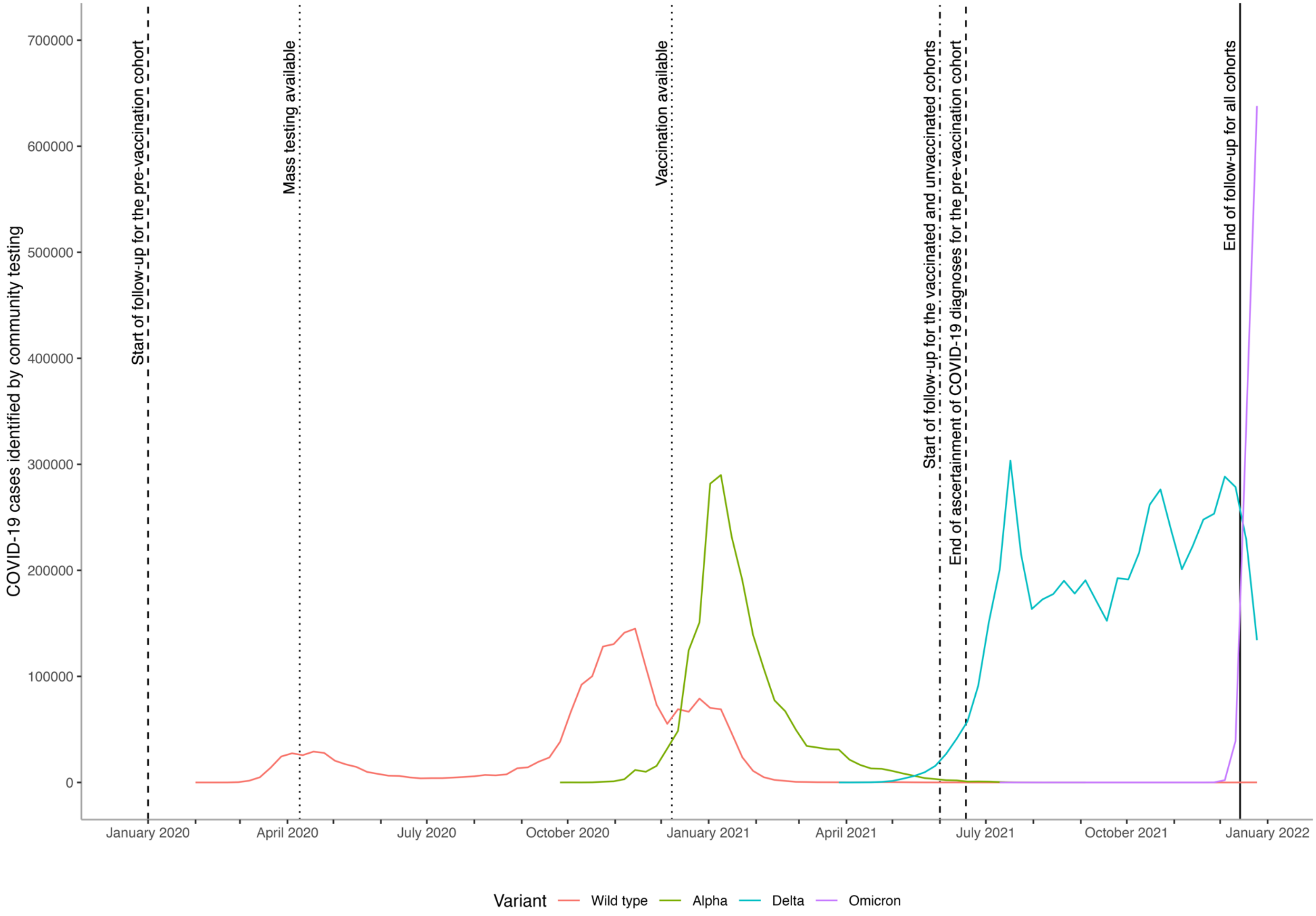

**Supplementary Figure 2: Diagram showing cohort construction.**

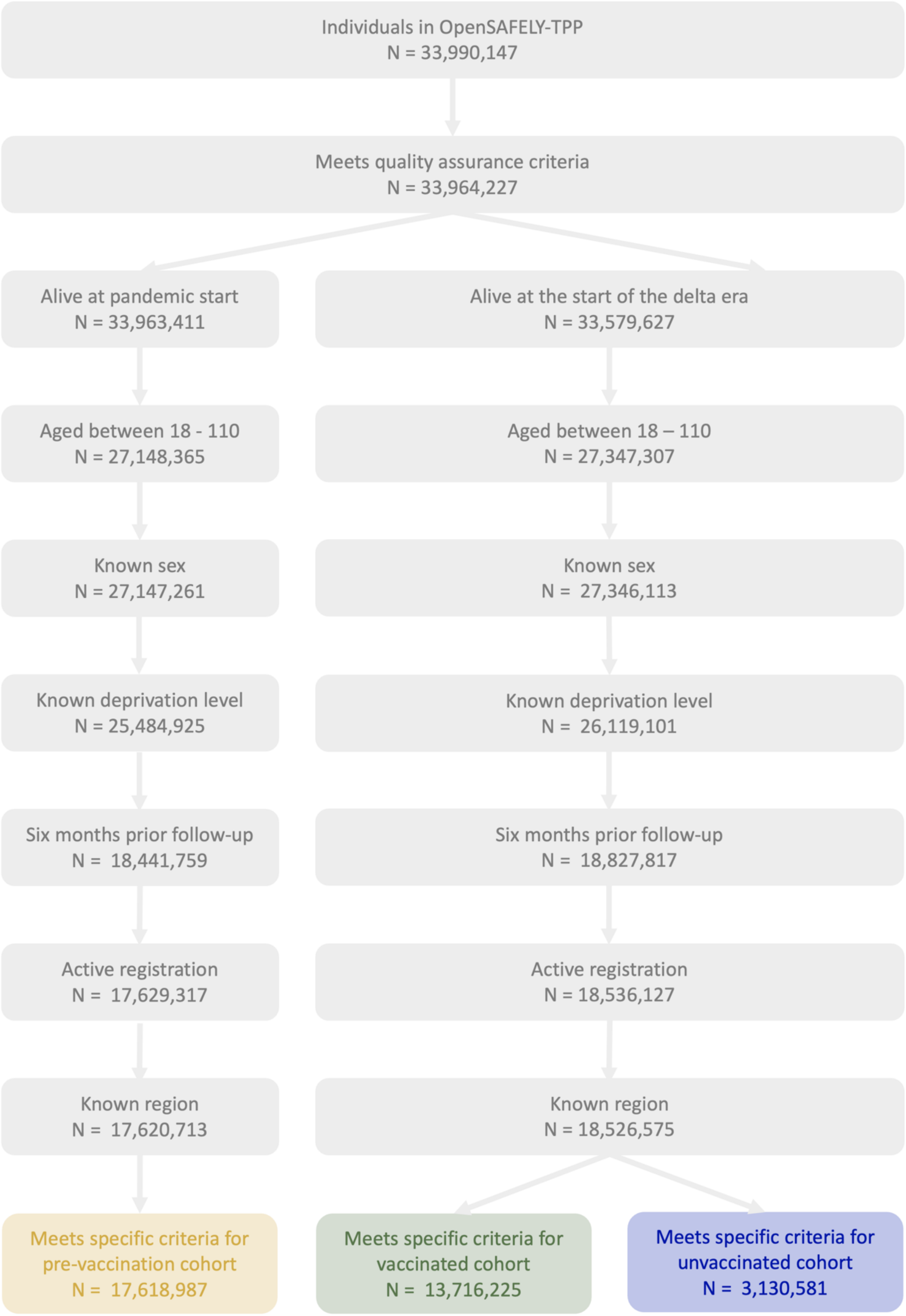

**Supplementary Figure 3: Maximally and minimally adjusted hazard ratios and 95% CIs for mental illness events following diagnosis of COVID-19 in the pre-vaccination, vaccinated and unvaccinated cohorts. Events on the day of COVID-19 diagnosis (day 0) were excluded.**

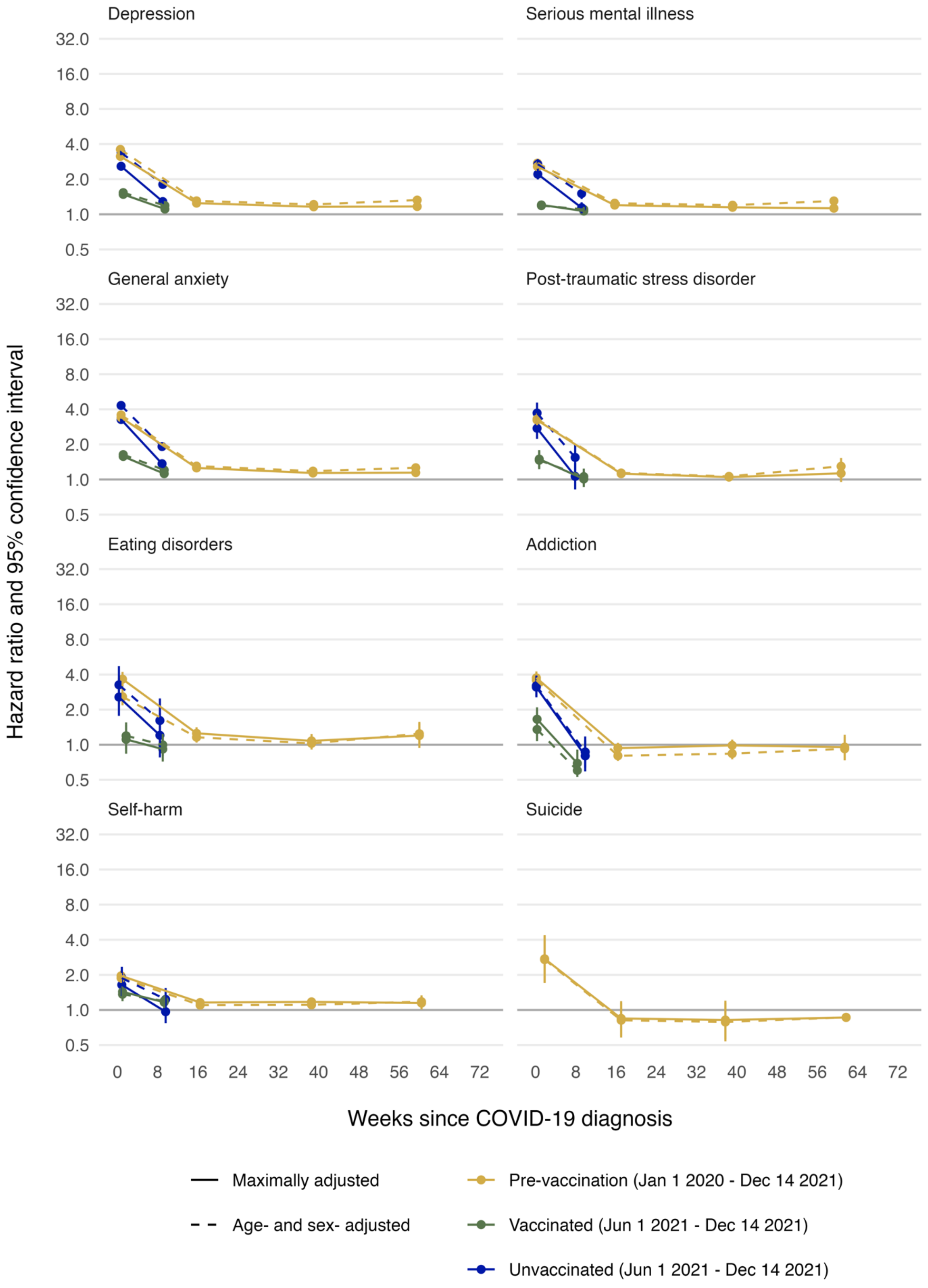

**Supplementary Figure 4: Maximally adjusted hazard ratios and 95% CIs for depression and serious mental illness following diagnosis of COVID-19 in the vaccinated and unvaccinated cohorts, by history of COVID-19. Events on the day of COVID-19 diagnosis (day 0) were excluded.**

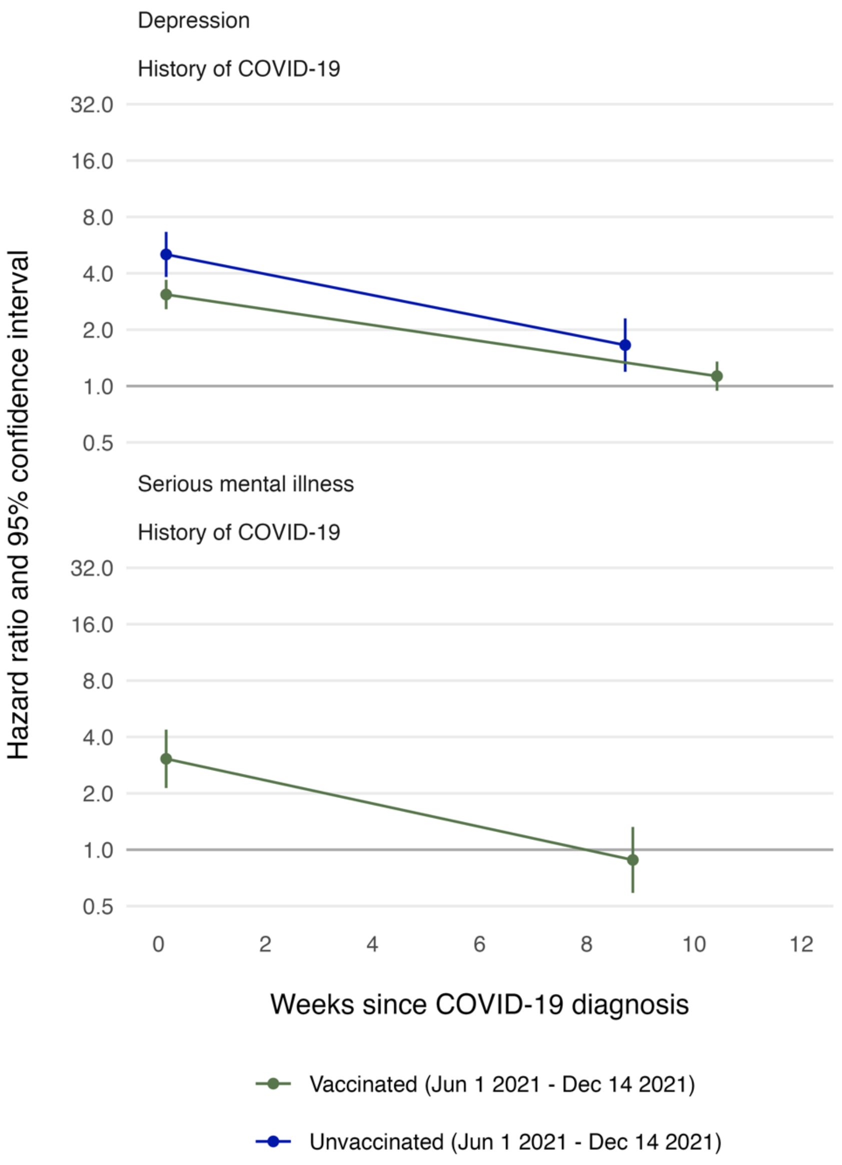

Note: there were less than 50 serious mental illness events post-COVID-19 in the unvaccinated cohort and so the model was not run.

**Supplementary Figure 5: Maximally adjusted hazard ratios and 95% CIs for depression and serious mental illness following diagnosis of COVID-19 in the pre-vaccination, vaccinated and unvaccinated cohorts, by age group. Events on the day of COVID-19 diagnosis (day 0) were excluded.**

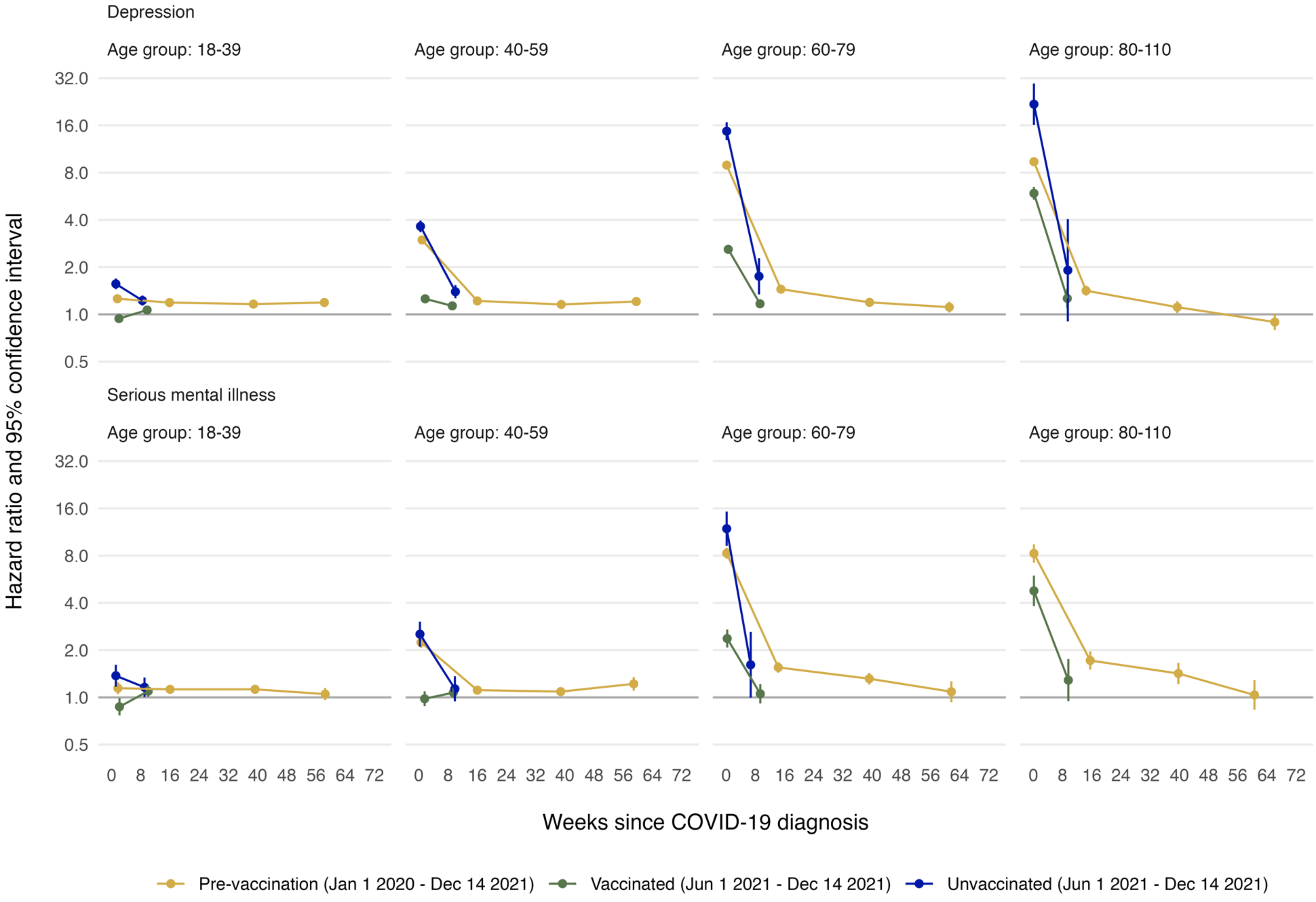

Note: There were less than 50 serious mental illness events post-COVID-19 among individuals aged 80-110 in the unvaccinated cohort and so the model was not run.

**Supplementary Figure 6: Maximally adjusted hazard ratios and 95% CIs for depression and serious mental illness following diagnosis of COVID-19 in the pre-vaccination, vaccinated and unvaccinated cohorts, by sex. Events on the day of COVID-19 diagnosis (day 0) were excluded.**

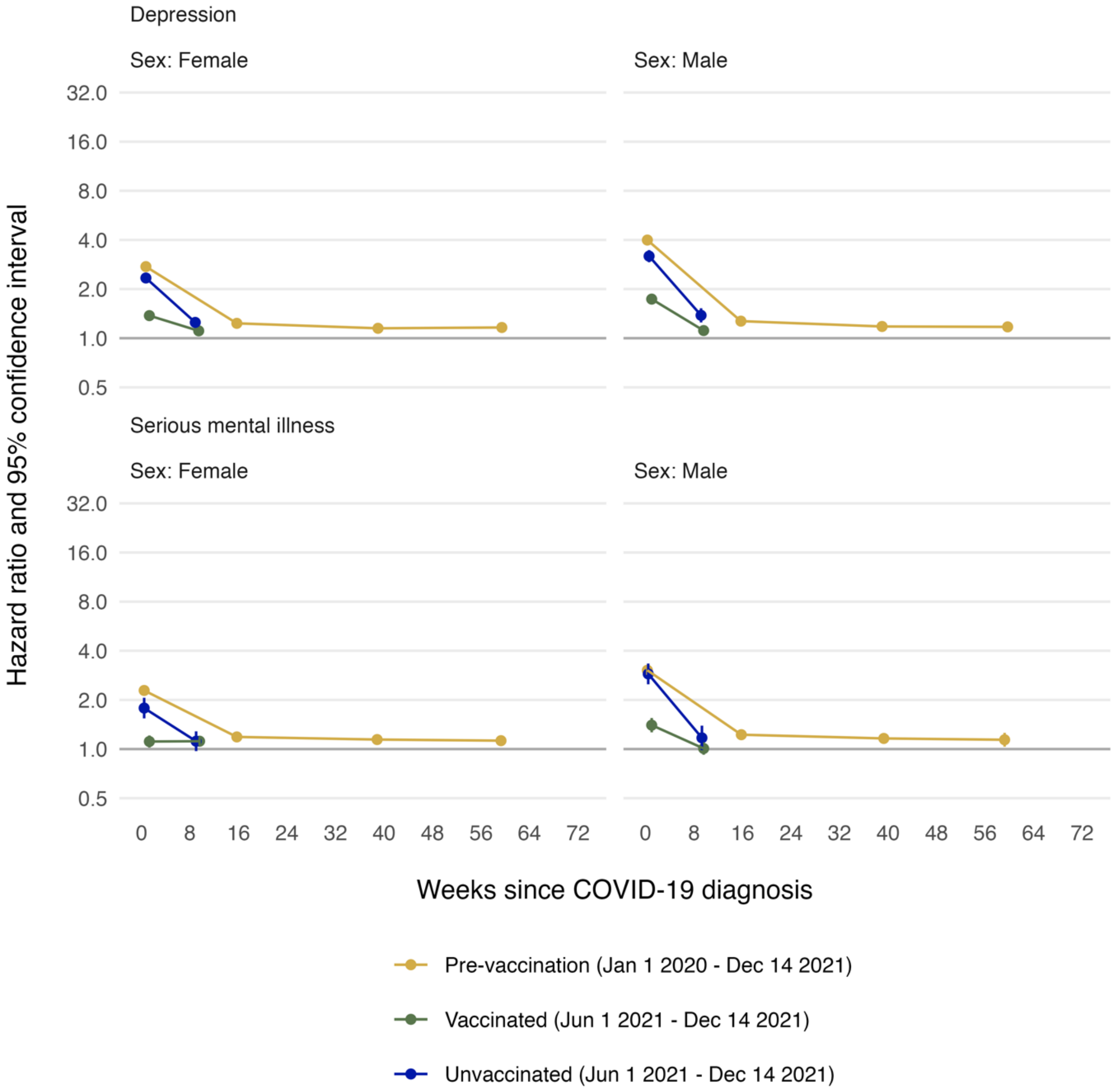

**Supplementary Figure 7: Maximally adjusted hazard ratios and 95% CIs for depression and serious mental illness following diagnosis of COVID-19 in the pre-vaccination, vaccinated and unvaccinated cohorts, by ethnicity. Events on the day of COVID-19 diagnosis (day 0) were excluded.**

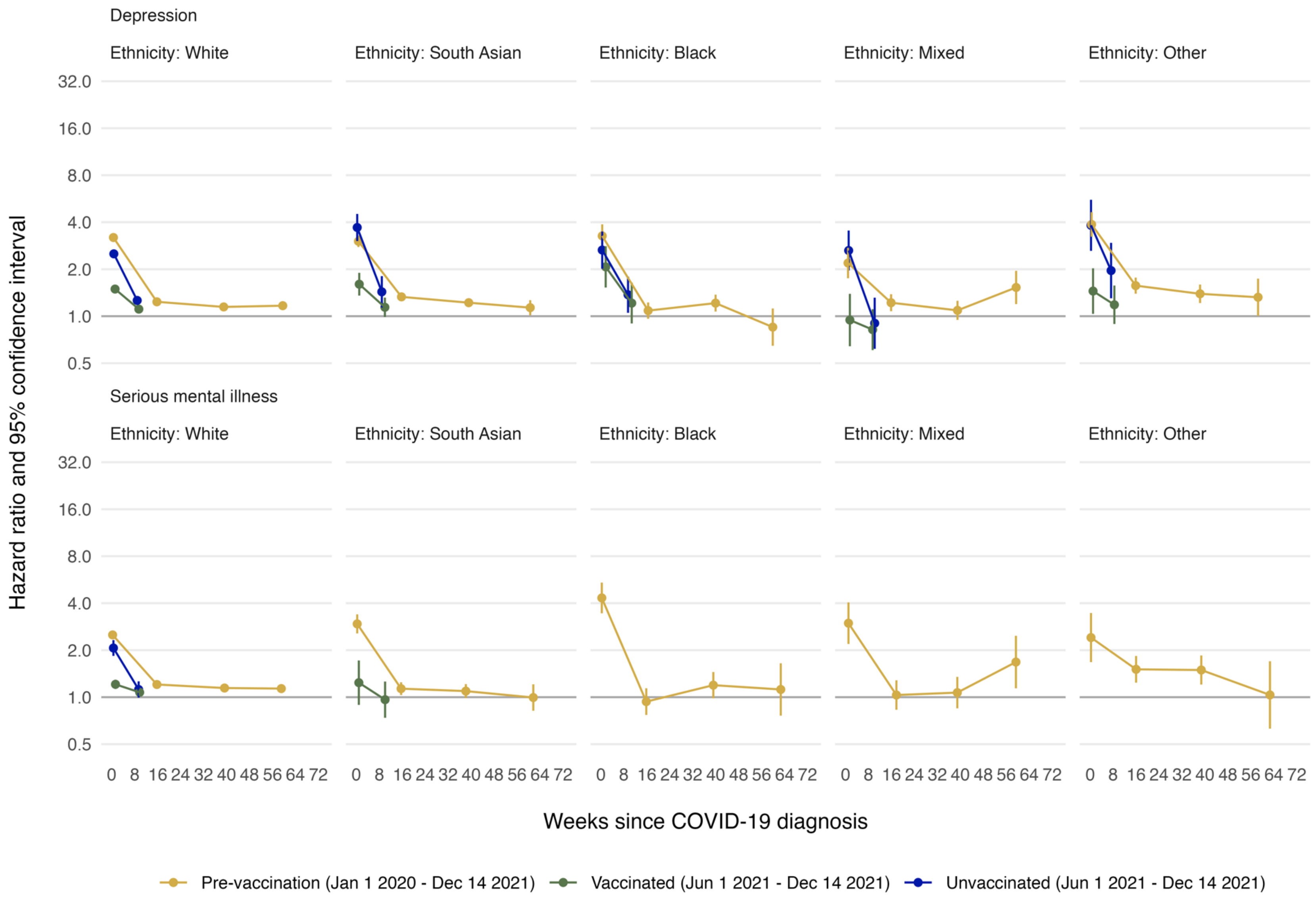

Note: There were less than 50 serious mental illness events post-COVID-19 among individuals of Black, Mixed, and Other ethnicities in the vaccinated and unvaccinated cohorts so the model was not run. There were also less than 50 serious mental illness events post-COVID-19 among individuals of South Asian ethnicity in the unvaccinated cohorts so the model was not run.
